# An ST131 clade and a Phylogroup A clade bearing a novel *Escherichia coli* O-antigen cluster predominate among bloodstream *E. coli* isolates from southwest Nigeria hospitals

**DOI:** 10.1101/2021.11.07.21265989

**Authors:** Ayorinde O. Afolayan, Aaron O. Aboderin, Anderson O. Oaikhena, Erkison Ewomazino Odih, Veronica O. Ogunleye, Adeyemi T. Adeyemo, Abolaji T. Adeyemo, Oyeniyi S. Bejide, Anthony Underwood, Silvia Argimón, Monica Abrudan, Abiodun Egwuenu, Chikwe Ihekweazu, David M. Aanensen, Iruka N. Okeke

## Abstract

*Escherichia coli* bloodstream infections are typically attributed to a limited number of lineages that carry virulence factors associated with invasion and, in recent years, invasive *E. coli* are increasingly multiply antimicrobial resistant. In Nigeria, *E. coli* is a common cause of bloodstream infections but the identity of circulating clones is largely unknown and surveillance of their antimicrobial resistance has been limited. We verified and whole genome-sequenced 68 bloodstream *E. coli* isolates recovered between 2016 and 2018 at three sentinel sites in southwestern Nigeria and susceptibility tested 67 of them. Resistance to antimicrobials commonly used in Nigeria was high, with 67(100%), 62 (92.5%), 53 (79%) and 37(55%) showing resistance to trimethoprim, ampicillin, ciprofloxacin and aminoglycosides, respectively. All the isolates were susceptible to carbapenems and colistin. The strain set included isolates from globally disseminated high risk clones including those belonging to ST12 (n=2), ST131 (n=12) and ST648 (n = 4). Twenty-three (33.82%) of the isolates clustered within two clades. The first of these consisted of ST131 strains, comprised of O16:H5 and O25:H4 sub-lineages. The second was an ST10-ST167 complex clade comprised of strains carrying capsular genes that may have originated in *Klebsiella*. We additionally determined that four temporally-associated ST90 strains from one sentinel were closely related enough to suggest that at least some of them represented a retrospectively detected outbreak cluster. Our data demonstrate that a broad repertoire of invasive *E. coli* isolates cause bloodstream infections in southwest Nigeria. In addition to pandemic lineages, particularly ST131, these include a previously undescribed lineage carrying an O-antigen cluster previously only reported from *Klebsiella*. Genomic surveillance is valuable for tracking these and other clones and for outbreak identification.

**Data Summary:** Phylogenetic tree, clinical data, and epidemiological data were visualized using Microreact (https://microreact.org/project/hmj3KwxS1dmmFPCKFx6qeA-invasive-escherichia-coli-sw-nigeria-2016-2018). All the sequence data have been deposited in the ENA under the project ID PRJEB29739 (https://www.ebi.ac.uk/ena/browser/view/PRJEB29739). Accessions can be found in Table S6.

## Introduction

Extra-intestinal pathogenic *Escherichia coli* (ExPEC) are responsible for the majority of infections in the blood, urine, meninges, prostate, and other normally sterile sites [1–3]. Although typically initially acquired within the gastrointestinal tract (and less commonly via genital route), these strains of *E. coli* differ from commensal and diarrhoegenic *E. coli* in possession of factors associated with systemic virulence [4, 5], allowing them to survive in different extra-intestinal niches. Some of the virulence genes associated with ExPEC include adhesins (*fim*, *pap*, *sfa*, *afa*), invasins (*ibeA*), iron acquisition genes (*ybt*, *iro*, *iuc*), toxins (*clb*, *cnf*, *hly*, SPATE genes) and protectins (*traT*, *ompT*, *kpsMT*), among others [4, 5]. ExPEC also often have K, O and H antigens that make them recognizable to *E. coli* experts and, in some instances, assist them in evading immune responses. Virulence and AMR determinants, as well as negative frequency-dependent selection, likely influence the stability of the most dominant ExPEC groups, which are often multiply resistant [6], thereby sustaining the occurrence of extra-intestinal diseases globally. In Africa, available data reveal that there is an upward trend in the prevalence of globally dominant ExPEC lineages in humans [7] and animals [8], painting a grim picture for disease and antimicrobial resistance.

Robust surveillance is urgently needed to tackle antimicrobial resistance in a more robust and consistent manner within each country. Whole genome sequencing (WGS) has helped to provide a clearer picture on the epidemiology of infectious diseases caused by ExPEC and has identified a number of pandemic clones. Incorporation of WGS with existing epidemiological frameworks in national public health institutes is critical for providing genomic context for prospective surveillance and for designing and implementing AMR-eliminating strategies.

Although studies conducted in Africa and other low- and middle-income countries (LMICs) have shown the abundance of invasive *E. coli* [9, 10], these studies are too few and far between so that ExPEC and AMR epidemiology are poorly understood [7, 11]. In Nigeria, there is sparse molecular information on ExPEC, but a few studies point to likely clonal expansion of resistant lineages and local presence of pandemic clones of concern [12–16]. These studies provide valuable information but represent an insufficient picture of ExPEC clones in Nigeria with few data available from the South. To extend the body of knowledge on genomic epidemiology of ExPEC in south western Nigeria, including AMR patterns and mechanisms, we leveraged on our existing genomic surveillance of bacterial AMR efforts by characterizing the genomes of invasive isolates from tertiary hospitals in South-west Nigeria.

## Methodology

### Species Validation and Antimicrobial Susceptibility Tests

Sentinel hospital laboratories referred anonymized clinical data and bloodstream *E. coli* isolates collected between the years 2016 to 2018 to the National Reference Laboratory for verification of isolate identity and antimicrobial susceptibility tests (AST). Isolate identity and AST were achieved using the VITEK2 Instrument. Drugs tested include amikacin (AMK), gentamicin (GEN), ampicillin (AMP), amoxicillin/clavulanic acid (AMC), piperacillin/tazobactam (TZP), cefuroxime (CXM), cefuroxime axetil (CXMA), cefepime (FEP), ceftriaxone (CRO), cefoperazone/sulbactam (SFP), nitrofurantoin (NIT), nalidixic acid (NAL), ciprofloxacin (CIP), ertapenem (ERT), imipenem (IPM), meropenem (MEM), and trimethoprim-sulfamethoxazole (SXT). Resistance profiles were generated from VITEK2 AST data. Antibiotic susceptibility results were interpreted in line with the CLSI 2019 standards [17]. Minimum Inhibitory Concentration values were converted to RIS values and the bug-drug combination table was generated using the AMR R package (v1.4.0; https://github.com/msberends/AMR) [18].

### Biofilm Assay

Following Wakimoto’s procedure [19], we sub-cultured invasive *E. coli* strains overnight in LB broth at 37°C with shaking at 160 rpm. Afterwards, we measured 190 µL of Dulbecco’s Modified Eagle’s Medium (DMEM) containing 0.45 % glucose using a pipette into each well of a 96-well plate (different plates were used for each time point (3h, 6h, 12h, 24 h)). Five µL of each isolate was inoculated in triplicate per time point into a 96-well plate from the stock plate and incubated at 37°C without shaking until each time point was reached. Absorbance at 595 nm was taken on completion of time point.

We pipetted spent media out of the 96-well plates. Each well was washed with PBS three times, fixed (10 mins, 200 µL of 75% ethanol), dried, and stained with 195 µL of 0.5 % crystal violet for 5 minutes. This was followed by washing and drying of the plates. We added 200 µL of 95% ethanol to each well, allowed the wells to stand for 20 minutes at room temperature, and determined absorbance with an enzyme linked immunosorbent assay (ELISA) plate reader at 570 nm. Biofilm index was defined using the average of the values for the optical density (OD) at 570 nm and 595 nm, and was calculated by dividing the OD values for each strain at a given timepoint by the OD values of the negative control at the given timepoint [20]. Enteroaggregative *E. coli* strain 042 was used as positive control while *E. coli* K-12 DH5α was used as negative control. The magnitude of biofilm formed by different genetically defined subgroups of isolates was compared using a two-tailed Mann-Whitney test at p<0.05.

### Haemolysis Test

Overnight LB cultures of the isolates were spotted onto the surface of the blood agar. Alpha haemolysis and beta haemolysis is indicated by a green colouration and a clear zone around bacteria colonies, respectively. Uropathogenic *E. coli* isolate ATCC 11175 and *stx*-deleted *E. coli* O157 isolate ATCC 7000728 were used as controls.

### DNA Extraction, Library Preparation, and Whole Genome Sequencing

Genomic DNA was extracted following the protocol outlined in a previous report [21]. Briefly, the Wizard DNA extraction kit (Promega; Wisconsin, USA; Cat. No: A1125) was used in the extraction of genomic DNA of all isolates. A dsDNA Broad Range quantification assay was used in the quantification of DNA extracts (Invitrogen; California, USA; Cat. No: Q32853). DNA libraries were prepared and sequenced using the NEBNext Ultra II FS DNA library kit (New England Biolabs, Massachusetts, USA; Cat. No: E6617L) and Illumina HiSeq X10 instrument (Illumina, CA, USA), respectively.

### Genome Assembly and Speciation

Raw sequence reads were assembled using the GHRU pipeline (https://gitlab.com/cgps/ghru/pipelines/assembly) which is summarily explained by the GHRU *de novo* assembly protocol [22]. Speciation was possible using Bactinspector (v0.1.3; https://gitlab.com/antunderwood/bactinspector/), implemented within the GHRU pipeline.

### Single Nucleotide Polymorphism (SNP) Analysis and Phylogenetic Tree Generation

The complete chromosome sequence of *Escherichia coli* strain EC958 (accession GCF_000285655.3) (https://www.ncbi.nlm.nih.gov/assembly/GCF_000285655.3/) was used to infer a whole-genome alignment of the sequence reads and identify SNP positions, which were, in turn used to infer a maximum-likelihood phylogenetic tree as per the GHRU mapping-based phylogeny protocol [22] which summarizes the steps implemented within the GHRU SNP phylogeny pipeline (https://gitlab.com/cgps/ghru/pipelines/snp_phylogeny). Isolates that clustered closely within the phylogenetic tree, belonged to the same ST, and shared similar resistance profiles, plasmid profiles, geography and dates of isolation, were investigated further for clue on potential outbreaks. We selected the closest reference genome to the outbreak isolates using Bactinspector, aligned the outbreak isolates (ST90) to the reference genome of *E. coli* strain D3 (accession NZ_CP010140.1; https://www.ncbi.nlm.nih.gov/assembly/GCF_001900635.1/), used Gubbins [23] to mask recombinant regions, and calculated pairwise SNP distances from the pseudo-genome alignment using FastaDist (v0.0.7; https://gitlab.com/antunderwood/fastadist), SNP-dists (v0.8.2; https://github.com/tseemann/snp-dists), and/or the R package harrietr (v0.2.4; https://github.com/andersgs/harrietr). The adegenet R package (v2.1.5; https://github.com/thibautjombart/adegenet/) was used to generate a pseudogenome alignment in DNA.bin format (one of the input files accepted by harrietr) from a pseudogenome alignment file. Phylogenetic tree, epidemiological data and in silico data were visualized in the web-based viewers Microreact [24], and the interactive Tree Of Life (iTOL) [25].

### AMR gene, Virulence gene, and Plasmid Replicon Prediction

The program SRST2 (v0.2.0; https://github.com/katholt/srst2/) [26] and the ARGannot database (https://raw.githubusercontent.com/katholt/srst2/master/data/ARGannot_r3.fasta) were used to screen raw sequence reads for the presence of acquired resistance genes. We also validated the report/output by utilizing the GHRU AMR Pipeline (https://gitlab.com/cgps/ghru/pipelines/dsl2/pipelines/amr_prediction) as explained in this protocol [22]. *ampC1* and *ampC2* were excluded from downstream analysis and visualization as they are beta-lactamase genes present in almost all *E. coli* isolates and are unlikely to confer antibiotic resistance in *E. coli* [27]. Overall plot of resistance determinants was constructed using the *upset* function from the UpSetR package (v1.4.0) [28]. Plots of AMR genes stratified by sequence type and sentinel site were constructed using the ggupset package (v0.3.0). Association between virulence genes and phylogroups of *E. coli* was determined using the *fisher_test* function from the rstatix package (v0.7.0). The calculation is based on single genes (including those that make up an operon), not operons. The associations were corrected for multiple testing using the Bonferroni method offered by rstatix. Bar plots were visualized using the *ggplot* function from the Tidyverse package (v1.3.1) in R.

The raw reads were screened for virulence genes with the GHRU pipeline, which utilizes ARIBA [29] and the VFDB database [30]. Plasmid replicon types were determined with same GHRU pipeline but using the PlasmidFinder database instead [31].

### Multilocus Sequence Typing (MLST) profiling

Following the Achtman MLST scheme [32], multi-locus sequence types (STs) were determined using the SRST2 program. We confirmed the results by using the GHRU MLST pipeline, which also implements the Achtman scheme. as summarized in the aforementioned GHRU protocol [22].

### *In silico* Serotyping and Phylogroup Determination

The O and H serotypes of *Escherichia coli* were determined using the SRST2 program and the EcOH database (https://raw.githubusercontent.com/katholt/srst2/master/data/EcOH.fasta). This result was validated using ECTyper (v1.0.0; https://github.com/phac-nml/ecoli_serotyping) and SerotypeFinder (https://bitbucket.org/genomicepidemiology/serotypefinder/src/master/). SerotypeFinder utilizes KMA [33] and BLAST+ [34] to predict *E. coli* serotypes from the alignment of raw and assembled reads, respectively, against the SerotypeFinder database. *Escherichia coli* genomes were assigned into phylogroups using ClermonTyping (v20.03) [35].

### Concordance

The agreement between phenotypic and genotypic antimicrobial resistance was determined for beta-lactams, cephalosporins, amikacin/kanamycin/gentamicin, trimethoprim, and the quinolones. Metrics such as sensitivity, specificity, true positives, true negatives, false positives, and false negatives were determined using the R script (https://gitlab.com/-/snippets/2050300, first used in a previous report) which utilizes the *epi.tests* function within the epiR package (v2.0.26) for each antimicrobial tested. Here, “Sensitive” and “Intermediate” values were combined, taking cognizance of the arguments for and against the use of the term “Intermediate” in clinical settings [36].

### Genome Annotation, Comparative Genomics, and gene location prediction

Functional annotation of Onovel32 clade genomes was performed using Bakta v1.0.4 [37]. Genomes were compared and visualized using Artemis v18.1.0 [38], Artemis Comparison Tool v18.1.0 [39], and Clinker v0.0.21 [40]. The mlplasmid web tool [41] was used to predict whether clinically-relevant virulence genes were borne on plasmids or on chromosomes.

## Results

### Epidemiology and Species Identification

Three hospital laboratories in South-west Nigeria submitted retrospective bloodstream *Escherichia coli* isolates with clinical and epidemiological data between the years 2016 to 2018. Available data showed that isolates were collected from patients aged 1 day to 71 years, with 17 (25%) of the isolates recovered from children under 60 days old (range 1-20 days, median 9 days), who would be characterized as neonates. The isolates were submitted from the University College Hospital (UCH; n = 22), Obafemi Awolowo University (OAU) Teaching Hospitals Complex (n = 18), and Osun State University Teaching Hospital, Teaching Hospital (until recently known as Ladoke Akintola University) (LAU; n = 28).

Of the 68 invasive isolates confirmed as *E. coli* by whole-genome sequencing (WGS), 48 (70.6%) and 64 (94.1%) were correctly identified as *E. coli* by the sentinel biochemical testing and reference laboratory VITEK2, respectively. *Escherichia coli* isolates were often misidentified as *Klebsiella pneumoniae* (n = 8) or *Citrobacter freundii* (n = 6) at the sentinel laboratories, while the VITEK2 system misidentified *E. coli* as *Klebsiella pneumoniae* (n = 3) or *Enterobacter aerogenes* (n = 1).

### Phylogroups, serotypes, and sequence types of *E. coli* bloodstream isolates

*E. coli* sent from all three hospital sentinel laboratories spanned all *E. coli* phylogroups, with 18, 15, 19, 8, 2, 1, and 5 *E. coli* genomes classified within phylogroups A, B1, B2, C, D, E, and F, respectively. The most common STs among the 33 identified include; ST131 (n = 12), ST156 (n = 5), and 4 each of ST10, ST167 ST410, ST648, and ST90. These 7 STs accounted for 54% of the *E. coli* isolates. Of these, only STs 131 and 167 were found across the 3 sentinel sites (Figure 1, Table S4). Diverse lineages were recovered from all three sentinel sites (OAU = 10 STs; LAU = 18 STs, UCH = 15 STs) (Table 1). While ST131 genomes made up 63% of genomes within phylogroup B2 (the second most common phylogroup), ST10 and ST167 genomes accounted for 44% of genomes within phylogroup A (the most common phylogroup).

**Figure 1:**
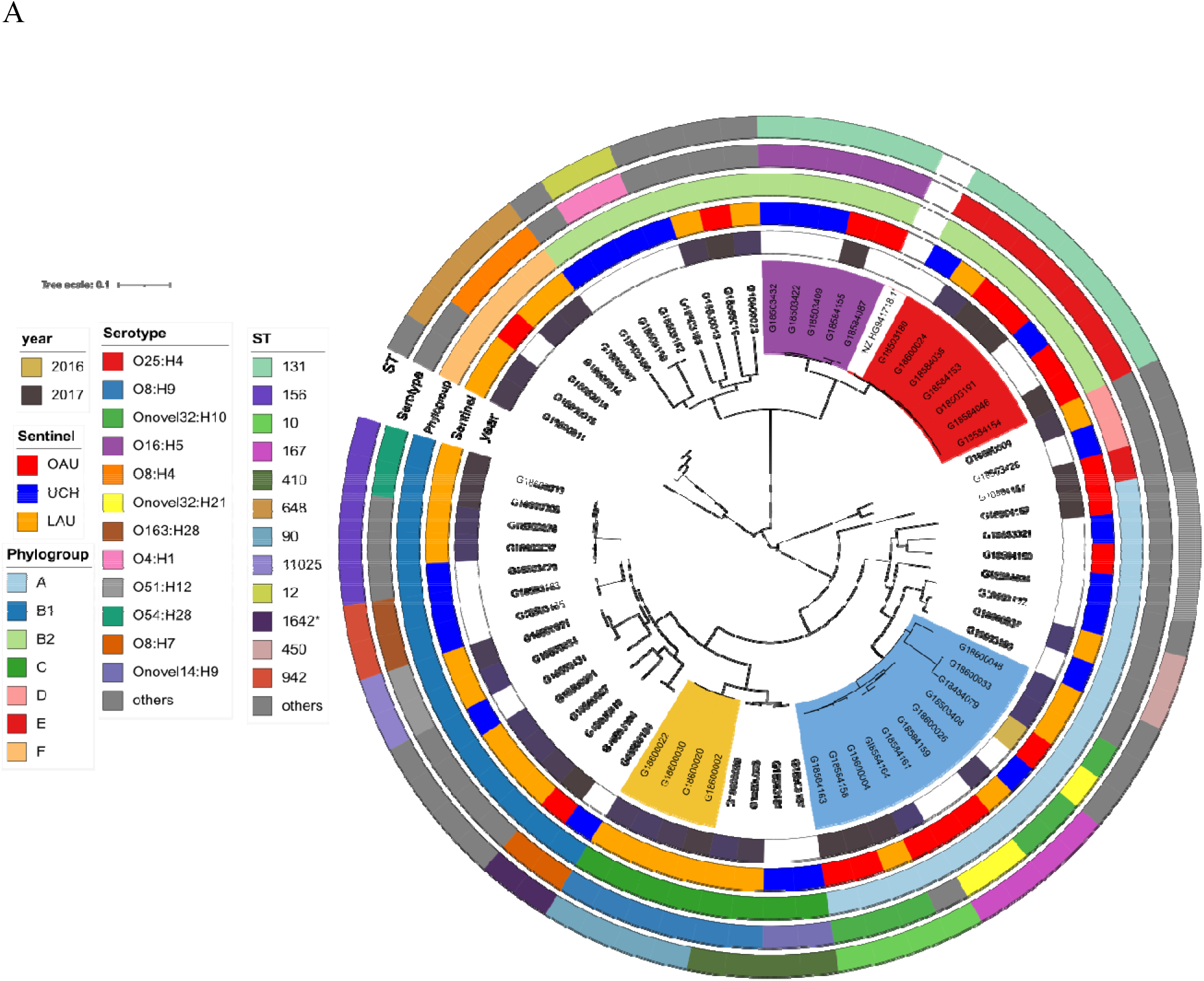
Maximum likelihood SNP tree of bloodstream *E. coli* isolates sequenced for this study. The purple-coloured clade represent ST131 lineage 1 (O16:H5 serotype); the red clade represent the ST131 lineage 2 (O25:H4 serotype); the blue clade represent the ST10-ST167 clade; The light brown clade mark the cluster from a single site representing likely outbreak clone of ST90 (b) The ST90 likely outbreak cluster of O8:H9 strains, showing resistance and plasmid profiles

**Table 1:**
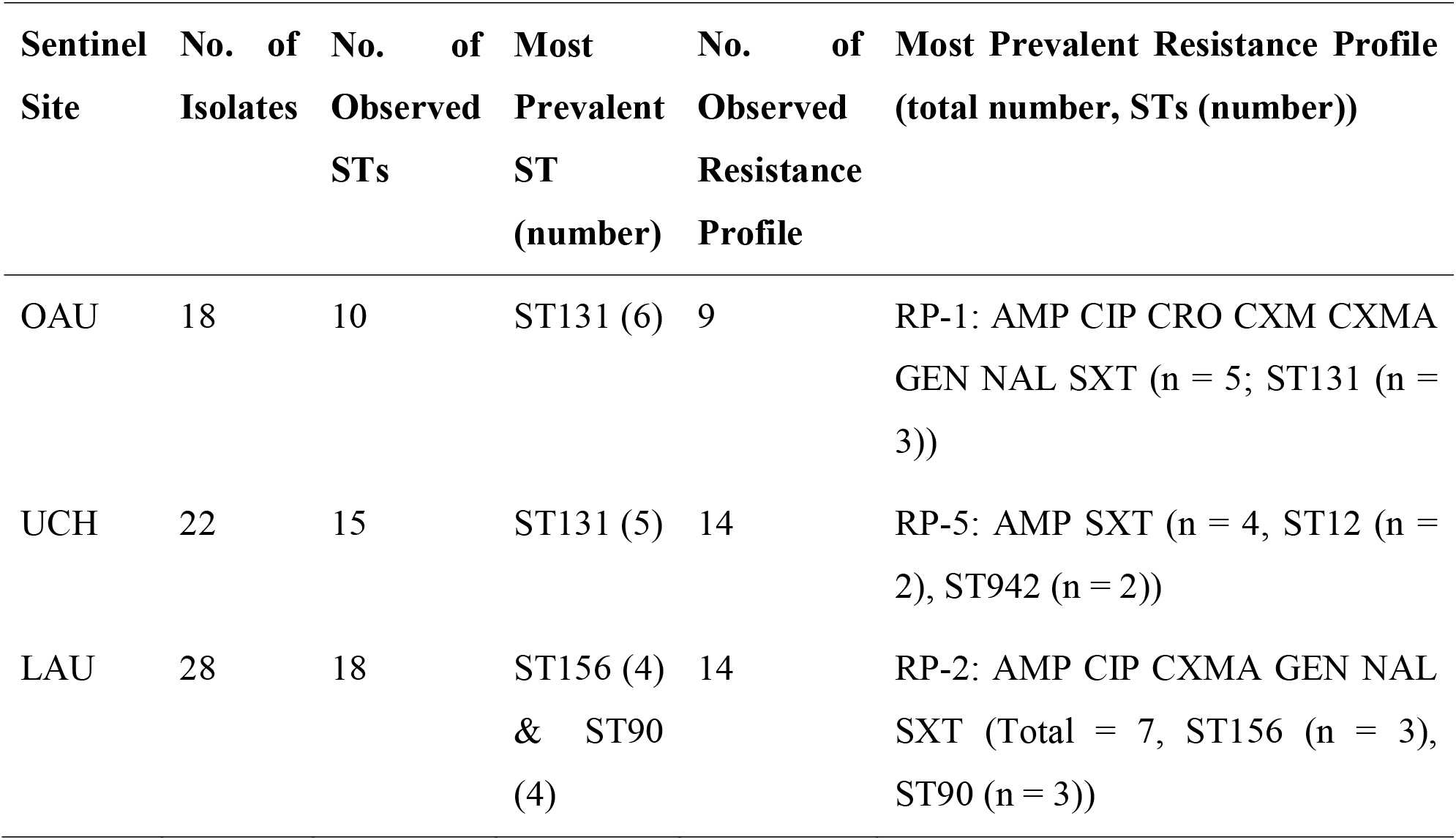
Distribution of Sequence Types (ST) and Resistance Profiles (RP) within Sentinel Sites.

While none of the isolates were submitted as suspected outbreak strains,, four isolates recovered from LAU within one month (January 2017) were typed as ST90, serotype O8:H9, carried the same plasmid profile (IncFI; IncFIA; IncFIB_AP001918; IncQ1), AMR gene profile, and similar but not identical resistance profile (Table S4). These four isolates were resistant to trimethoprim, the quinolones, gentamicin, cefuroxime axetil, and ampicillin (Table S3b). Comparison of pairwise SNP differences revealed that two of the four isolates were identical while the other two isolates differed from these by 11 SNPs and 72 SNPs. The current literature supports a clustering threshold between the range of 0 to 17 SNPs for suspected outbreak-related *E. coli* blood isolates [42–44]. Although these isolates were correctly identified within this sentinel site, the cluster was only recognized retrospectively, likely due to different beta lactam resistance profiles.

ST131 was the most common sequence type detected and the 12 isolates belonging to this ST clustered into two distinct lineages defined by serotype H5 (n = 5; O16:H5) and the serotype H4 lineage (n = 7; O25:H4), and henceforth referred to as the ST131 lineage 1 and ST131 lineage 2, respectively (Figure 1).

A total of 38 unique serotypes and 25 O-groups were identified. Of note was the most common O-type, which was novel and identified in eleven isolates belonging to phylogroup A. These ONovel:32 strains belonged to ST10 or ST167, or were single or double locus variants of these STs. They included six ONovel32:H10 isolates as well as three H21 and one H4 –flagellin-encoding strains. Along with one ONT:H10 strain that also belonged to ST10, they formed a distinct clade on the phylogenetic tree (Figure 1). Strains belonging to this cluster were submitted from all three hospitals. Two of them were originally misclassified as *Klebsiella pneumoniae* by VITEK2 at the reference laboratory level. Irrespective of whether the ST90 clade is discounted, the ONovel:32 clade and the ST131 clade were the most abundant. Together they accounted for 32.4 % of the isolates and both clades were found in all three hospitals (Figure 1).

### Virulence Factor Profiles of the Bloodstream *E. coli* Isolates

Diverse virulence genes were observed among the ExPEC genomes: 159 virulence-associated (VAG) genes were found at least once in the 68 bloodstream isolate genomes. Enterobactin genes (*entB*, *entC*, *entE_1*, *entS*) and ferrienterobactin precursors and proteins (*fep* operon genes (ABCDEG) and *fes_1*), were found in more than 95% of the isolates. The outer membrane hemin receptor (*chu*), siderophores (*fyuA*, *irp*, *ybt*), intimin-like adhesin (*fdeC*), haemolysin (*hly*), aerobactin (*iuc*), polysialic acid transport protein *kpsM_1*, pyelonephritis-associated pili *pap*, SPATE genes (*sat*, *vat*), and plasmid-encoded *Shigella* enterotoxin *senB* were more abundant and significantly more associated with phylogroup B2 than with phylogroups A, B1, and C (Fisher’s exact Test, p < 0.05; Figure 2a). Figure 2b shows that a wide range of biofilm-forming capacities were seen in the subset of isolates tested with moderate or strong biofilm-formers being most common in phylogroups B1 and B2 as well as the ST90 outbreak strain-containing phylogroup C. No significant difference in biofilm-formation between the phylogroups was observed.

**Fig. 2:**
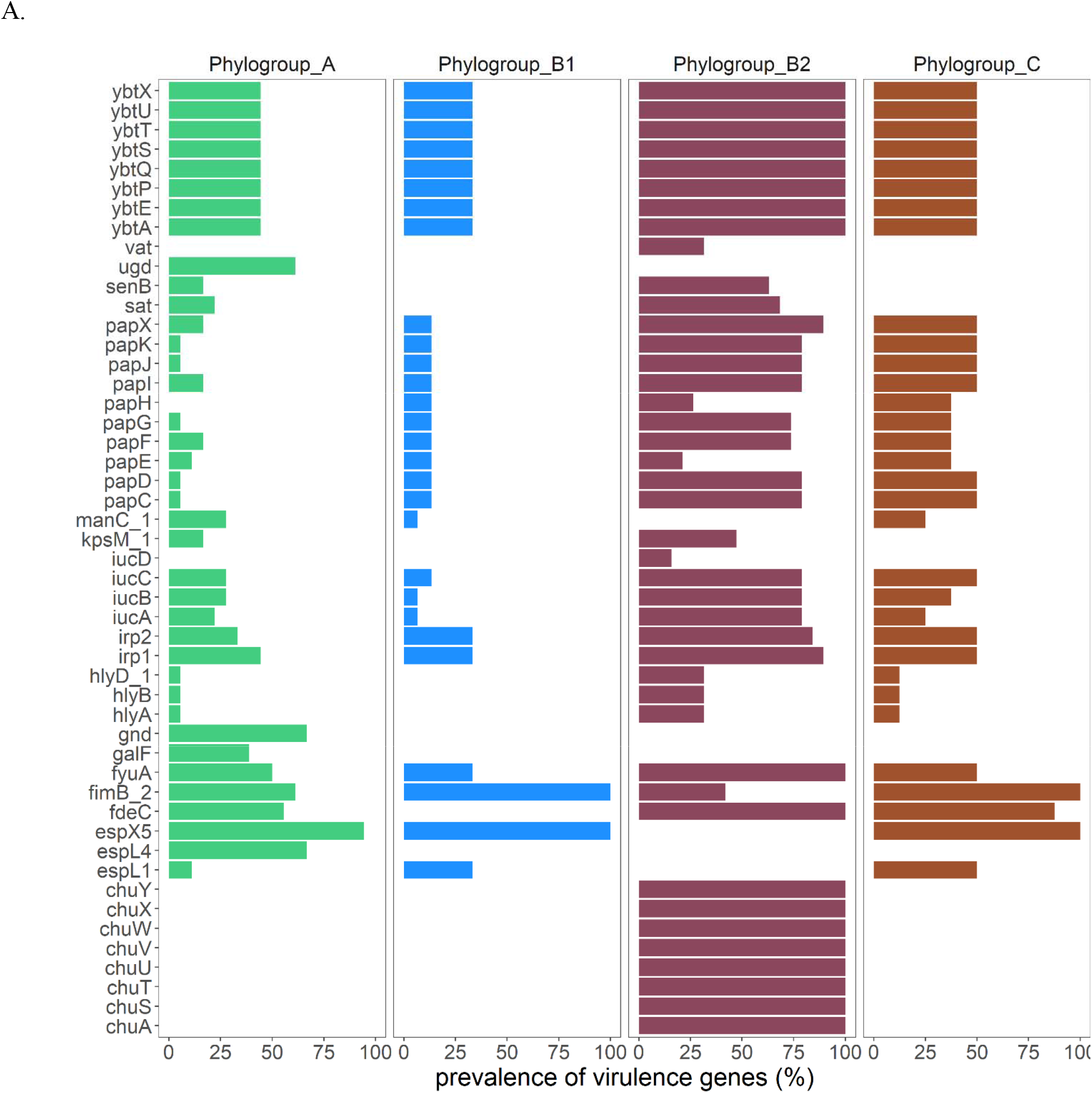
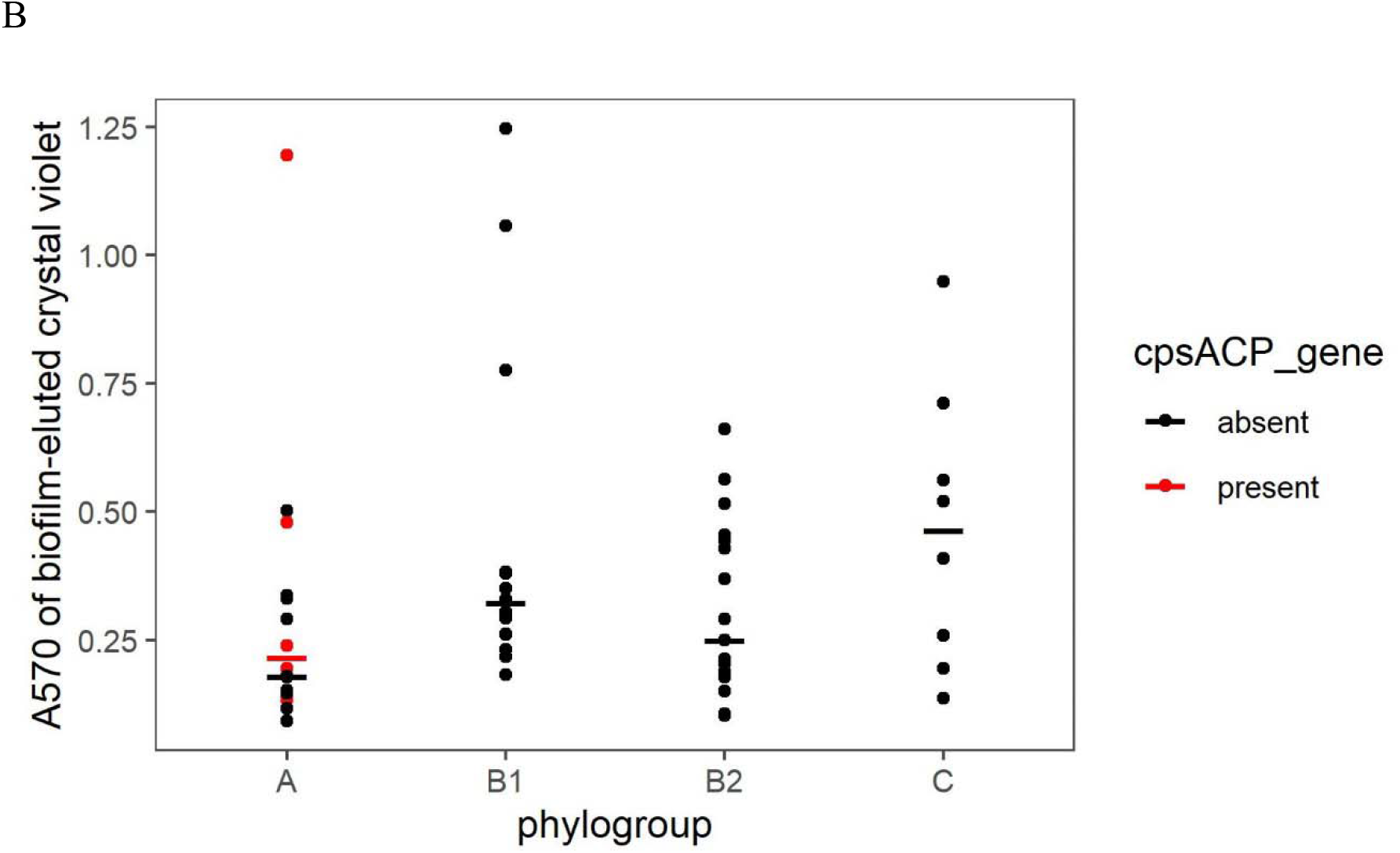
a. Comparison of virulence genes among isolates clustered within phylogroups A-C. Graph shows the prevalence of virulence factors in isolates within phylogroups containing more than five isolates: A (n = 18), phylogroup B1 (n = 15), phylogroup B2 (n = 19), and phylogroup C (n = 8). Using Fisher’s test (p < 0.05), only the VAGs significantly more prevalent in at least one phylogroup are shown. B. Biofilm formation in 67 strains, measured as A570 nm of crystal violet eluted from fixed and stained six-hour biofilms. Each dot represents data from a single strain belonging to the phylogroup listed on the X axis. Horizontal bars mark the median for each phylogroup, outliers inclusive due to the small number of tested strains in each phylogroup. The A570 nm values for isolates carrying the cpsACP gene were represented as red dots.

Phylogroup B2 isolates, comprised largely of ST131 strains, carried the highest number of virulence-associated genes (n = 86 VAGs). Thirty-four VAGs were significantly more common in ST131 isolates (n = 12) than in non-ST131 isolates (n = 56). Of these the genes encoding the outer membrane haem receptor (*chu*), yersiniabactin (*ybt*), and pyelonephritis-associated pili (*pap*) were found in more than 84% of the ST131 isolates (Fisher’s exact Test, p < 0.05; Fig. 3a). On the other hand, 6 VAGs were significantly more common in non-ST131 isolates than in ST131 isolates, including 2 genes (*gspK* and *gspL*) present in more than 85% of the non-ST131 isolates, but in only about 50% of ST131 isolates. The VAGs *entD*, *espL1*, *espX1,* and *espL5* were absent in ST131 isolates but were present in at least 32% of non-ST131 isolates, notably Phylogroup A ONovel32 strains (Fig. 3a).

**Fig. 3.**
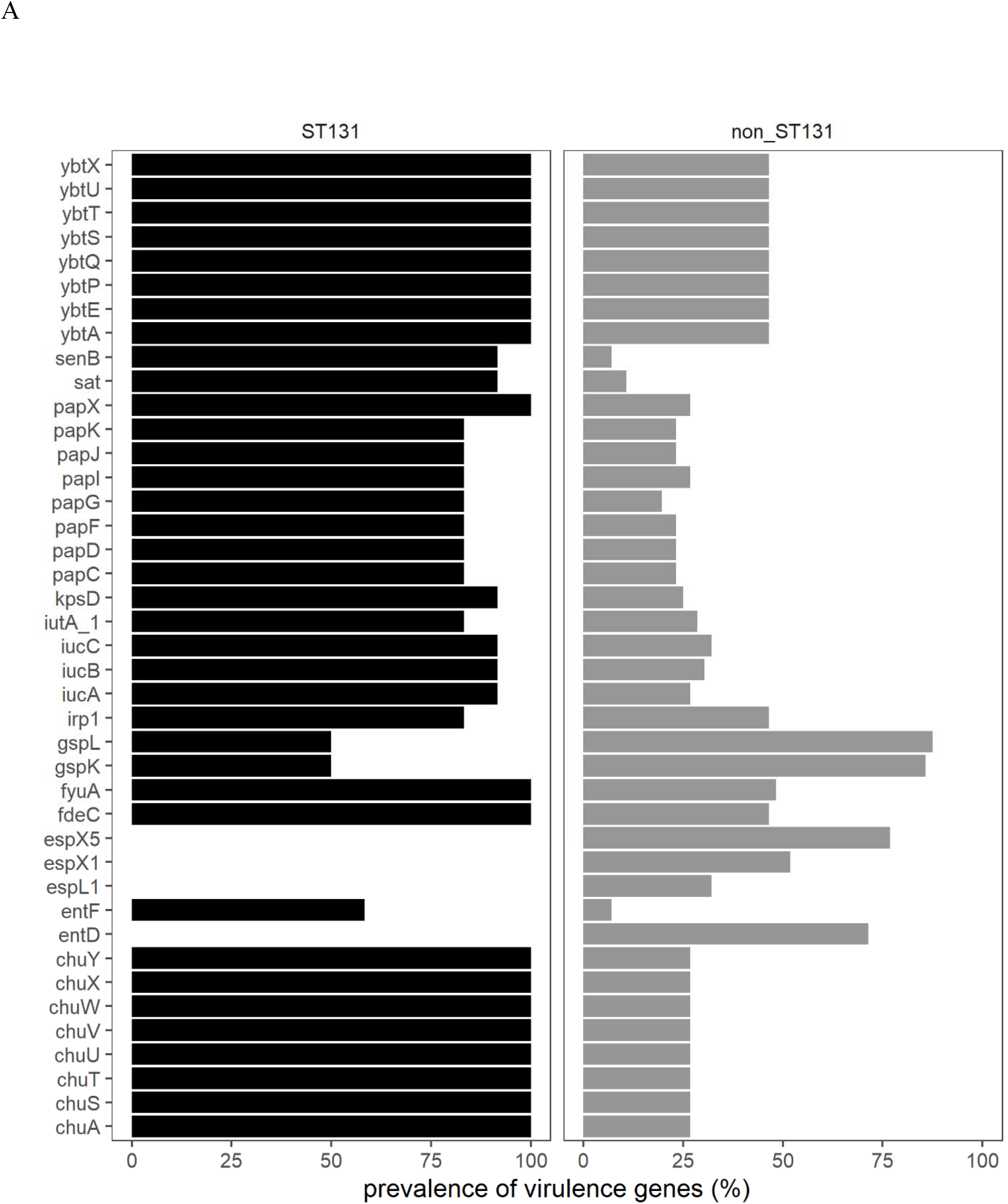
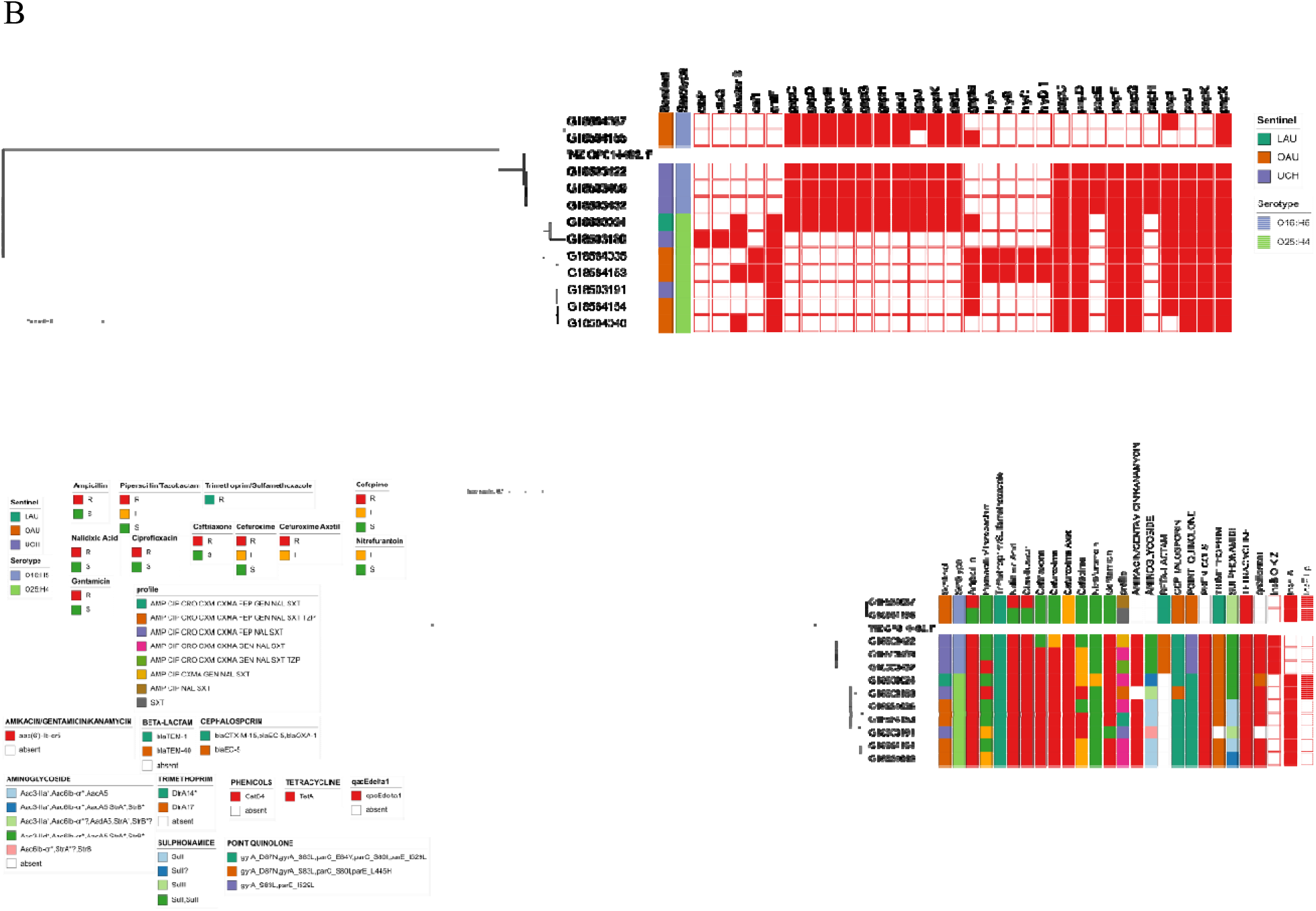
a: Comparison of virulence genes between ST131 (n = 12) and non-ST131 (n = 56) isolates. Using Fisher’s test (p < 0.05, Bonferroni-corrected), only the VAGs significantly more prevalent in the STs are shown. b: Virulence factor profile of isolates belonging to ST131. The presence or absence of a virulence gene is indicated by a red or white colour on the heatmap, respectively. c. Antimicrobial resistance (AMR) phenotype profile and AMR genotype profile of isolate belonging to ST131. The presence or absence of plasmid Inc types is indicated by a red or white colour on the heatmap, respectively.

Both lineages of ST131 share common VAGs, including; adhesins, yersiniabactin, aerobactin, enterotoxin, and transport-associated genes but some genes were only seen, or predominantly seen in one of the two lineages. For example, the haemolysin (*hly*) carried by two isolates within ST131 lineage 2 were absent in ST131 lineage 1. Also, two isolates within ST131 lineage 2 carried the cytotoxic necrotizing factor *cnf1*. (Figure 3a and 3b). The median pairwise distance between isolates in ST131 lineage 1 is 2857.5 SNPs (range: 0 – 2866), while the median pairwise distance between isolates within ST131 lineage 2 is 499 SNPs (range: 0 – 827). The inter-clade SNP distance is 13619 (range: 12900 – 14146) (Figure S1).

None of the ONovel32 clade isolates carried alpha haemolysin or pyelonephritis associated pili, which were present in ST131 and some of the other lineages. However, some virulence-associated genes were predominant among or restricted to ONovel32 strains (Figure 4a and 4b). These include the *esp4L* type III secretion effector. We also searched independently for type III secretion systems and identified a cluster 97.5% identical to *E. coli* type III secretion system 2, previously associated with virulence in septicemic *E. coli* [45], in all *esp4L*-positive and one *esp4L*-negative Onovel32 isolate (GI8584164). *esp4L* was more common among these isolates than other phylogroup A strains as well as multiple genes encoding capsular modification enzymes. All the ONovel32 strains and one associated ONT strain carried a *ugd* gene. The only other isolates in this collection with this gene were three isolates belonging to the globally disseminated high risk clone ST648 [46], known to possess biofilm-associated features that enhance pathogen emergence and persistence in both the human body and the environment. The *ugd* gene is associated with hypermucoviscosity and invasive virulence and the ONovel32 allele is 96% identical to that from *K. pneumoniae* NTUH-K2044, a hypervirulent *K. pneumoniae* isolate [47] and *K. variicola* (Accession number CP079802.1) capsular cluster *ugd* genes. Six of the ONovel:32 strains (but not the ONT:H10 strain in the same clade) carried *cpsACP*, a chromosomally-borne gene, which is predicted to encode a phosphatidic acid phosphatase (PAP2 Pfam 01569) family gene. PAP2 phosphatases replace phosphate groups on lipid A with amine groups resulting in a positively charged lipid A that confers resistance to cationic peptides [48]. PAP2 phosphatases have been known to be transmitted horizontally solitarily or as part of capsular clusters [49]. A BLAST search revealed that the ONovel32 PAP2 allele is 99.6% identical to endogenous PAP2 genes from *Klebsiella variicola* (Accession number CP079802.1). As shown in Figure 5, depicting the region for ONovel32 ST1284 strain OAU-VOA-056, *cpsACP* is located within a capsular gene cluster identical to a *K. variicola* cluster and flanked by a 5’ IS3 transposase and a 3’ IS1 protein InsB-encoding gene. At the opposite end of the cluster is the *ugd* gene. The cluster shows G+C content and other base-pattern signatures that depart from the *E. coli* flanking sequence (Figure 5a and 5b).

**Fig. 4.**
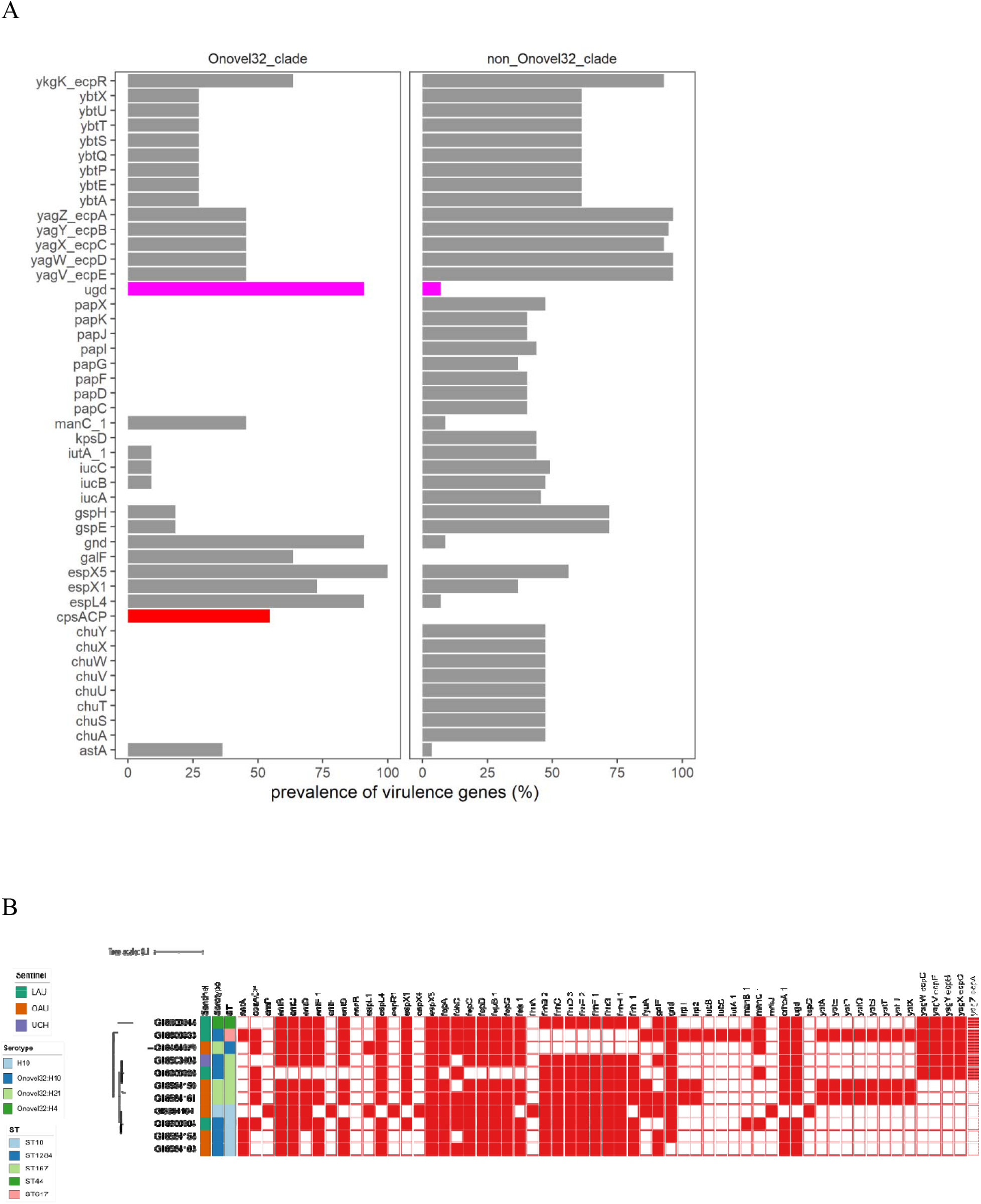
a: Comparison of virulence genes between Onovel32 (n = 11) and non-Onovel32 (n = 57) clade isolates. Using Fisher’s test (p < 0.05, Bonferroni-corrected), only the virulence genes significantly more prevalent are shown. The cpsACP gene and the ugd gene bars were coloured red and magenta, respectively. b: Onovel32 clade isolates belonging to the sequence types ST10, ST167, and their locus variants belong to phylogroup A. Leaf tip colours differentiate the sentinel sites; OAU (Red), UCH (Blue), LAU (Yellow).

**Figure 5.**
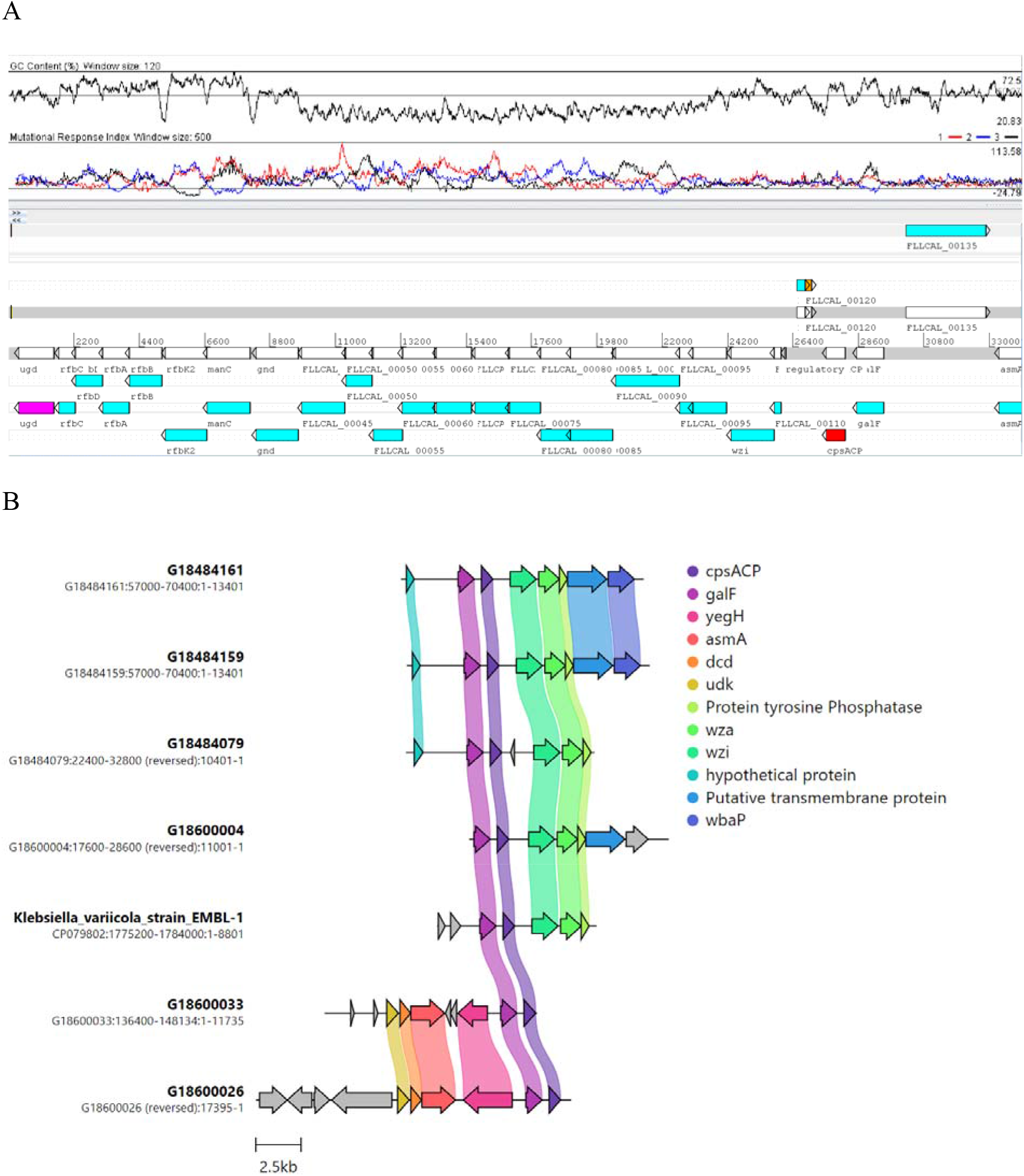
a: Schematic cluster in ST1294 ONovel32 chromosome that includes the *ugd* and *cpsACP* genes. The genes within the cluster are syntenic and identical with a cluster from *Klebsiella variicola* (Accession number: <u>CP079802.1</u>). Above the schematic depiction of gene is a G+C content plot and mutational response index plot. (b) comparison of the ONovel32 cluster with analogous cluster in *Klebsiella variicola* strain EMBL-1

Observing that ONovel:32 cluster isolates, some of which were originally misclassified as *Klebsiella*, showed mucoidity upon plate culture, we sought to determine whether these strains had distinctive colonization-associated capacities. The median A_570_ at six hours was 0.178 for phylogroup A strains lacking *cpsACP* and 0.216 for those with the gene (Figure 2), but these differences were not significant. All the isolates were also negative in the string test for hypermucoviscosity in *Klebsiella* [50].

### Resistance Profiles (RP) and Concordance with Predicted Antimicrobial Resistance

Susceptibility testing of 67 out of the 68 isolates showed that, of the 16 antibiotics tested, resistance to trimethoprim-sulfamethoxazole (n = 67; 100%), ampicillin (n = 62; 92.5%), nalidixic acid (n = 57; 85.1%), and ciprofloxacin (n = 53; 79.1%) was commonly observed (Fig. 6a, Fig. 6b, Table S2). On the other hand, resistance to cefoperazone.sulbactam (n = 3; 4.5%) was less common. All isolates were susceptible to amikacin, ertapenem,, and meropenem (Table S2, Fig. S1a and S1b). One *bla_CTX-M-15_* –positive isolate from LAU (ST11025, B1, O51:H12) was categorized as showing intermediate to imipenem but did not carry any carbapenemase-producing gene. Resistance profiles were remarkably similar among the three sentinels (Figure 6b). Resistance to trimethoprim/sulfamethoxazole among isolates belonging to all 33 STs could be explained by the possession of *dfrA* (n = 54/67) and *dfrB* (n = 2/67) genes. Ciprofloxacin resistance (23 STs) was largely attributable to mutation in the quinolone resistance determining regions (QRDR) of *gyrA* (D87N, S83L), *parC* (E84A, E84G, E84K, S57T, S80I), and/or *parE* (E460D, I355T, I529L, I529L, I464F, L416F, L445H, S458A, S458T), with or without the presence of plasmid quinolone resistance genes (*qnrS*, *qnrVC4*, *qepA*, *aac-(6’)-Ib-cr*). About 45% (n = 24/53) of isolates phenotypically resistant to ciprofloxacin carried a combination of *aac(6’)-lb-cr5* gene and mutation in the QRDR regions (*gyrA*, *parC*, and *parE*) (Figure 7a and 7b). The most common quinolone resistance gene profile observed is the “gyrA_D87N,gyrA_S83L,parC_S80I,parE_S458A”, as observed in a quarter (n = 14/53) of isolates showing phenotypic resistance to ciprofloxacin.

**Fig. 6.**
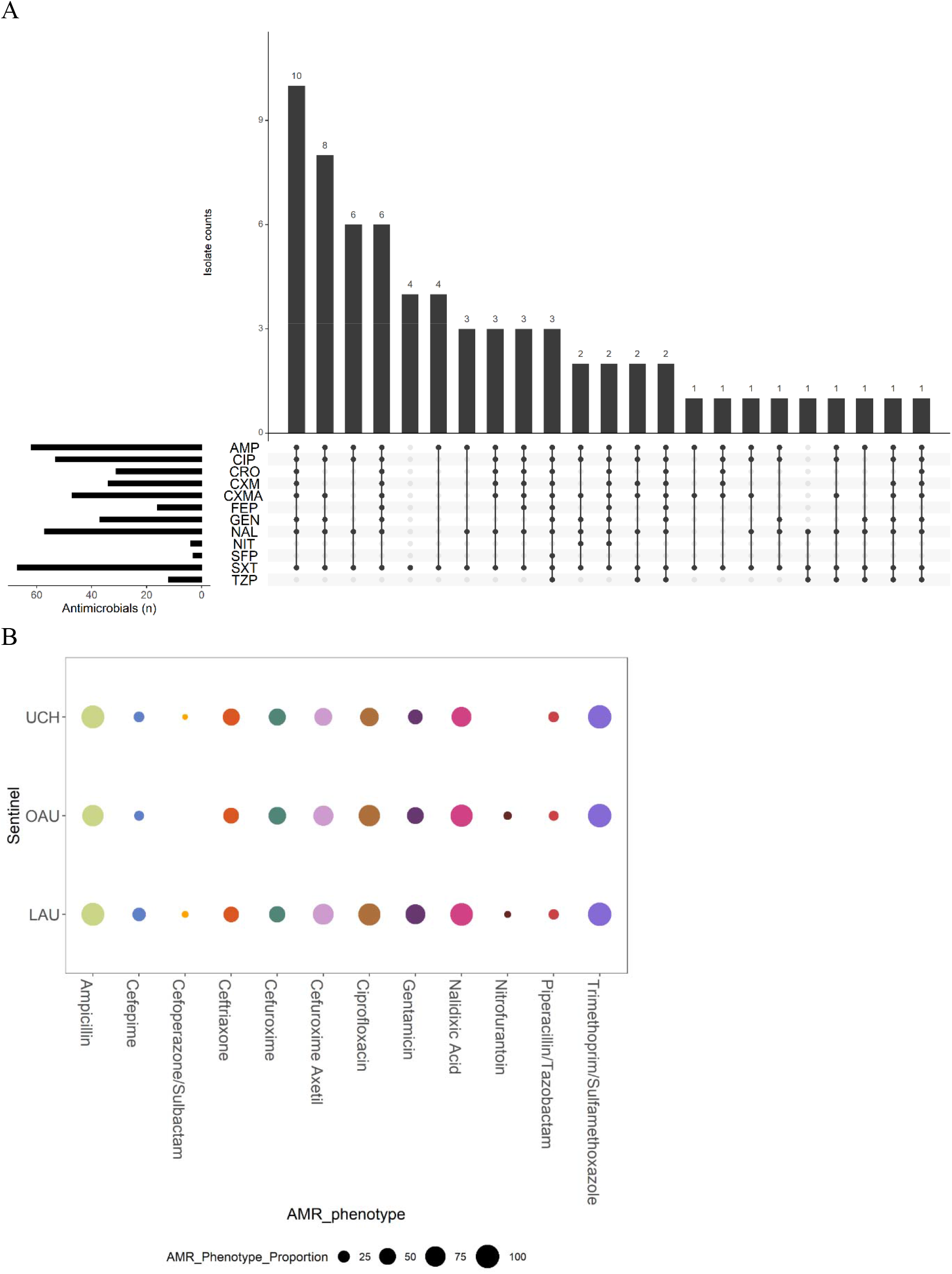
a: Resistance Profile of ExPEC isolates. The main bar chart demonstrates the number of ExPEC isolates with each combination of resistance to tested antibiotics, and is ordered in descending order by the frequency of resistance profiles observed among 67 ExPEC isolates. The side bar chart demonstrates the number of isolates that are resistant to each of the named antibiotics. The dots and lines between dots at the base of the main bar chart (and the right side of the side bar chart) show the co-resistance status of the ExPEC isolates. All isolates were susceptible to Amikacin, Meropenem, Imipenem, and Ertapenem. AMP = Ampicillin; CIP = Ciprofloxacin; CRO = Ceftriaxone; CXM = Cefuroxime; CXMA = Cefuroxime Axetil; FEP = Cefepime; GEN = Gentamicin; NAL = Nalidixic Acid; NIT = Nitrofurantoin; SFP = Cefoperazone/Sulbactam; SXT = Trimethoprim/Sulfamethoxazole; TZP = Piperacillin/Tazobactam Fig. 6b: AMR phenotypes of ExPEC Isolates, stratified by Sentinel Site (LAU (n = 28), OAU (n = 22), UCH (n = 21)). The size of the coloured circles represent the proportion of isolates recovered from each sentinel site that demonstrated resistance to the tested antibiotics.

**Fig. 7:**
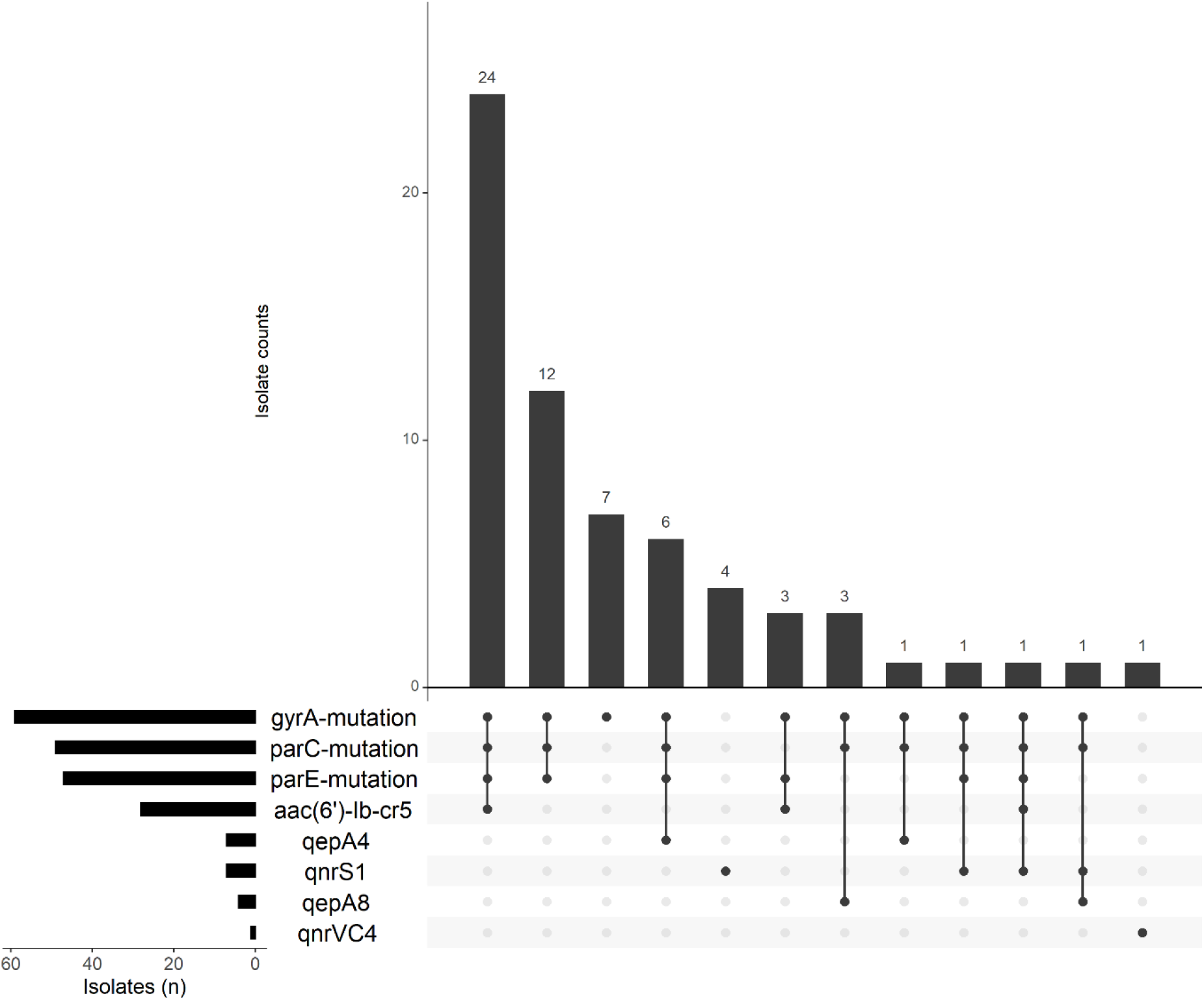
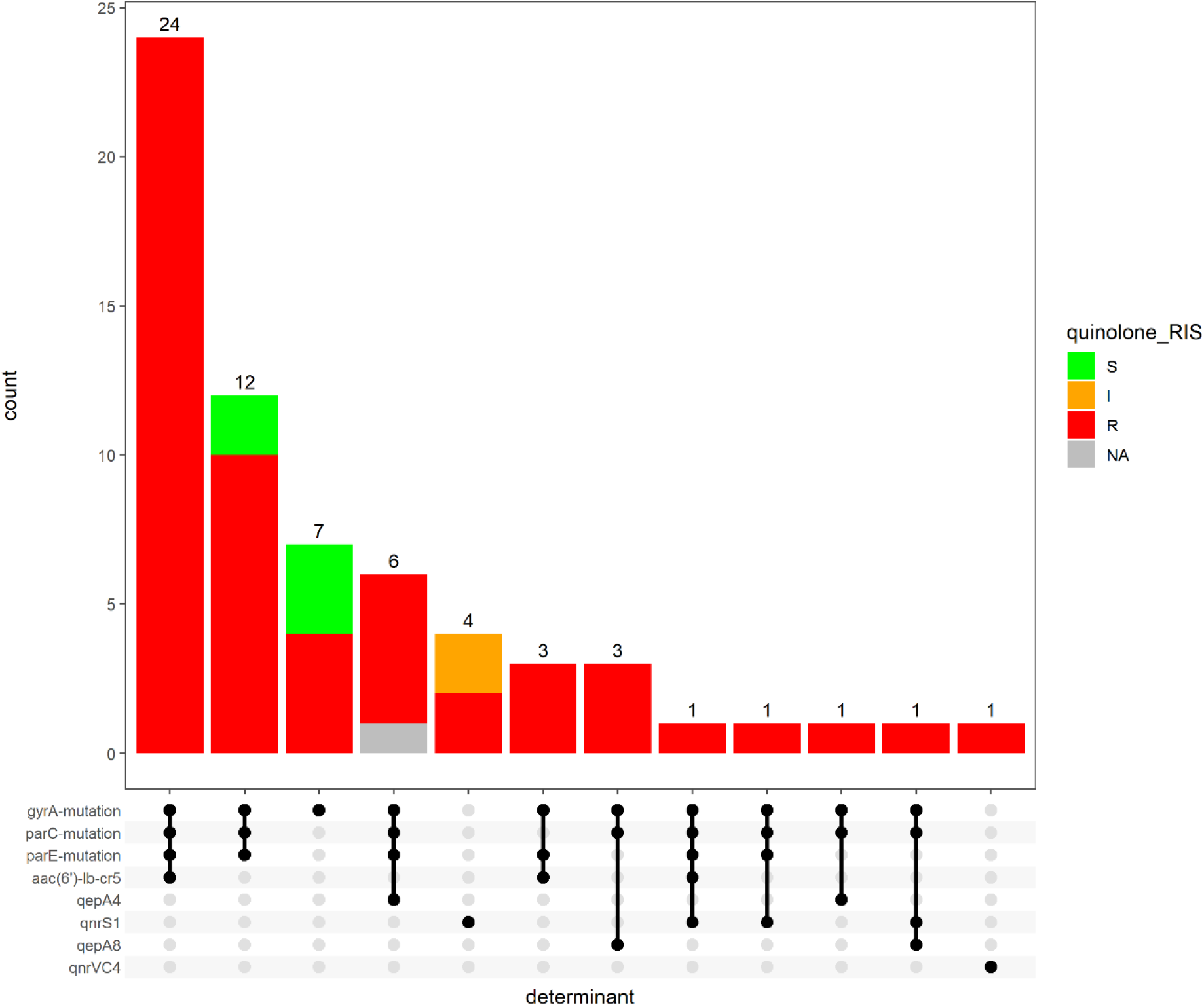
Quinolone resistance gene combinations in ExPEC isolates. The upset plots show (a, b) the number of ExPEC isolates carrying each combination of genes conferring resistance to the quinolones (b), and is coloured by the proportion of observed phenotypic antimicrobial susceptibility, and is ordered in descending order by the frequency of resistance gene profiles observed. The side bar chart demonstrates the number of isolates that carry each of the resistance genes. The dots and lines between dots at the base of the main bar chart (and the right side of the side bar chart) show the co-resistance gene profile of the ExPEC isolates. From the legend of the coloured upset plot, NA means that phenotypic antimicrobial susceptibility data for the isolate is absent.

Ampicillin resistance among isolates belonging to 29 STs (n = 62) could be explained by the carriage of a range of beta-lactamase genes including *bla_TEM-1_* (36/62 of isolates), *bla_TEM-40_* (3 isolates), *bla_TEM-84_* (2 isolates), *bla_TEM-135_* (2 isolates),, or *bla_OXA-2_* (co-occurring with *bla_TEM-90_*; 1 isolate). Extended-spectrum beta-lactam resistance likely resulted from *bla_CTX-M-15_*, *bla_CTX-M-27_*, and *bla_CMY-42_* (Figure S2a and S2b) co-occurring largely with the plasmids Inc types FIB_AP001918, FIA, IncFI, IncQ1, IncFII_p, and the col plasmid Col156.

Of the 5 drug classes tested, the highest concordance between phenotypic resistance and predicted antimicrobial resistance was observed for trimethoprim (100% concordance, TP = 67/67, Sensitivity = 100%, Specificity = NA) and the lowest concordance for drugs within the aminoglycoside class (concordance = 55.22%, TP = 37/67, Sensitivity = 100%, Specificity = 0 %) (Table S3a).

### Multidrug Resistance (MDR)

We observed a total of 23 resistance profiles (RP) (Table S1) with resistance profiles in many cases associated with specific STs (Table 1 and Table S4). Fifty-nine (88.06%) of the isolates were resistant to at least one agent within at least three classes of antibiotics, fitting the multi-drug resistant (MDR) definition of the international AMR community [51] (Table S1). Isolates belonging to the most frequent sequence types form the bulk of invasive isolates carrying ESBL genes or mutations in the quinolone-resistance determining regions (*gyrA*, *parC*, *parE*), and the plasmid-mediated quinolone resistance gene *aac(6’)-lb-cr,* as well as genes conferring resistance to trimethoprim. Carbapenemase genes were conspicuously absent, in agreement with the AMR phenotype. Almost all isolates carrying ESBL genes (n = 32/34) are multidrug resistant. About 30% of these showed the most frequent resistance profile (RP-1) (Figure S3). Furthermore, multi-drug resistant bloodstream isolates do not carry the same resistance determinants and plasmid replicons, apart from *gyrA/parC/parE* mutations (Figure S3).

Resistance gene profiles differed significantly between the two ST131 clades (Figure 3c). Unlike ST131 lineage 1 isolates, ST131 lineage 2 isolates did not carry any beta-lactamase gene besides *ampC*. Within the ST131 lineage 1, two isolates did not carry genes conferring resistance/reduced susceptibility to the aminoglycosides, phenicols, macrolides, and quaternary ammonium compounds. Furthermore, the absence of the Col156, IncB_O_K_Z, and IncFI plasmids in these two isolates seem to have been compensated by the possession of IncFIA, IncFII_p, and IncI1 plasmids, noted to have been absent in the other three ST131 isolates within the ST131 lineage 1.

All the ONovel32 strains (but not the co-clustering ONT strain) carried one ESBL gene, *bla_CTX-M-15_*, and these strains also carried IncF plasmids, common among phylogroup A strains (Table S4, Table S5). Every one of them carried the most common four resistance-conferring mutations in the QRDRs (gyrA_D87N,gyrA_S83L,parC_S80I,parE_S458A) and seven, including the ONT strain, carried *qepA4.* Four ONovel32 isolates additionally carried *aac-(6’)-Ib-cr,* conferring aminoglycoside and ciprofloxacin resistance, as well as horizontally-transmitted genes conferring resistance to trimethoprim, chloramphenicol and tetracyclines were common (Microreact link, Table S5).

Common to both the ST131 and the ONovel:32 clades is the high prevalence of *qacEdelta1* (n = 9 and 8, respectively) conferring resistance to quaternary ammonium compounds, commonly used for disinfecting hospital surfaces and associated with class 1 integrons. Altogether, these clades comprised 17 (42.50%) of 40 *E. coli* isolates carrying this gene.

## Discussion

This study characterized 68 bloodstream *E. coli* isolates as an important first step in understanding their epidemiology within south-west Nigeria. In this small collection, we identified multiple clones of pandemic importance, and found two predominant clades. One of these, comprised of two ST131 lineages is globally disseminated and this study illustrates its importance in Nigerian health facilities. The second predominant clade does not feature in present discourse on international ExPEC clones and represents strains belonging to ST10, ST167 and related STs, which predominantly encode genes that could confer a novel O-antigen type. In addition to these prominent clades, we identified strains belonging to major pandemic ExPEC lineages, including ST12, ST73, and ST648, and their single locus variants. ST69, ST95, and ST405 lineages were not detected but our sample is not very large and therefore our findings are insufficient to rule them epidemiologically insignificant in our setting. We additionally identified in the collection five ST90/410 strains, all from a single facility. Two ST90 isolates are identical, differ from a third by only 11 SNPs, and likely represent an outbreak. Our findings add to information that is chronicling ExPEC lineages of importance within Nigeria [14–16, 52, 53], other parts of Africa [16, 54], and other low-middle-income countries [16, 55].

Among ST131, we found both globally-disseminated lineages within our isolate collection. The majority of haemolytic and *pap* gene-bearing phylogroup B2 strains belonged to this ST. Biofilm formation among these strains was common and associated with the *kpsD* gene, a known contributor to biofilm formation [56]. The ST10, ST167 and related strains that comprised the ONovel32 clade were distinguished by the presence of one of two variations of a capsular island that has been well described in *Klebsiella* with biofilm formation among them being more pronounced among strains carrying the *cpsACP*-containing portion of the island. While the virulence of ST131 has been well described, features of this clade that cause it to predominate remain unknown and further studies are required to understand its pathogenicity and selective success in our setting.

High levels of phenotypic resistance to antibiotics within the antifolate (trimethoprim. co-trimoxazole), quinolone (nalidixic acid), fluoroquinolone (ciprofloxacin), cephalosporin (cefuroxime axetil), and the aminoglycosides (gentamicin) classes observed among invasive isolates in this study corroborates reports from previous studies [12, 52, 53] and pose a serious concern for clinical therapeutics. *In silico* data confirm the abundance of genes conferring resistance, with many isolates carrying more than one gene conferring resistance to a specific antimicrobial or class. Before the turn of the millennium, resistance to many of the aforementioned antimicrobials was rare in Africa. However, steady increase in the availability and use of these agents in the empiric treatment of ExPEC-related infections has inevitably selected for AMR. The increased rate of fluoroquinolone resistance in diarrhoeagenic *E. coli* and other enterobacterales, for instance, coincided with increased fluoroquinolones use in Nigeria [57] and other parts of Africa [7, 58]. While findings from a study conducted in Nigeria more than a decade ago [13] concluded that nalidixic acid was still an effective antimicrobial, we observed in this study that resistance to nalidixic acid is now common.

Similarly, cephalosporin resistance emerged and expanded much more in Africa than in other parts of the world as these agents became the drug of choice for treating multi-drug resistant pathogens [58]. Fluoroquinolone resistance has also been previously associated with the presence of ESBL genes because ESBL genes are often borne on transferable large plasmids that co-host some of the PMQR genes [59]. We observed the co-carriage of ESBL and fluoroquinolone resistance genes in more than a quarter of the ExPEC isolates, and particularly in over-represented lineages.

We find that both predominant lineages we have highlighted in this report show multiple resistance. This has important implications for patients with life-threatening bloodstream infections and provides a plausible explanation for their evolutionary success in our setting. Resistance to antimicrobials used intensively in Nigeria (trimethoprim, aminopenicillins and ciprofloxacin) was rife and resistance to the agents most frequently used empirically when blood stream infections are suspected – second- and third-generation cephalosporins and aminoglycosides was also worryingly common. While none of these clades showed carbapenem or colistin resistance, these reserve antimicrobial classes are out of the reach of most patients attending the three hospitals from which the strains were obtained.

Next-generation sequencing has emerged as a promising complement to clinical bacteriology as it provides answers to medical conundrums as well as a more robust picture of the epidemiology of infectious diseases. It can also reveal, as in this case, circulation of hitherto unrecognized clones of concern. Although its integration into clinical diagnosis and patient care gaining ground many parts of the world, its adoption in LMICs (and the rest of the globe) is still hindered by lack of infrastructure, cost of implementing WGS, limited bioinformatics expertise and as yet mildly inaccurate prediction of antimicrobial resistance [60, 61]. In our study, we observed perfect concordance (100%) with phenotypic AST data for trimethoprim, but not for the cephalosporins, the quinolones, or the aminoglycosides. This therefore shows that further understanding of resistance mechanisms and routine AMR database update is needed to enhance the feasibility of gradual and sustained integration of WGS into routine diagnosis.

Another advantage of next-generation sequencing is its potential in rapid detection of outbreaks either retrospectively, or in real-time. We report a likely retrospective ST90 outbreak in LAU, which would not have been detected using traditional diagnostic methods in the sentinel laboratories. ST90 strains have variously been highlighted for their zoonotic potential and association with device-related hospital outbreaks [62–64] and its epidemiology in our setting remains to be understood ST90, and related ST410, were only seen at one facility and the representation of ST90 at this facility may have been amplified by an outbreak. The adoption of genomic surveillance in diagnostic laboratories within Nigeria will ensure that outbreaks of this and other clones can be detected in real time, while definite tracking and containment of the spread of such clones will be achieved before lives are lost.

This study has a few limitations. Blood culture is infrequently performed in Nigerian hospitals and until recently, most isolates were not archived. Therefore, these isolates represent but a fraction of the ExPEC likely to have infected patients in the three hospitals and may not be representative. Our short-read data makes it impossible to accurately determine whether the resistance genes are located on the bacterial chromosome or plasmids (or other mobile genetic elements). In future, we will incorporate long read sequencing into our prospective surveillance efforts in order to correctly identify plasmid-borne AMR genes.

In conclusion, this study provided hospital-specific information on the population structure of ExPEC lineages needed to track pandemic lineages and guide infection disease control practices in line with Nigeria’s national action plan on antimicrobial resistance.

## Data Availability

Phylogenetic tree, clinical data, and epidemiological data were visualized using Microreact (https://microreact.org/project/NigeriaEcoliInvasive/d0dc3660) and Phandango (for visualization of pangenome, tree, and metadata). All the sequence data have been deposited in the ENA under the project ID PRJEB29739 (https://www.ebi.ac.uk/ena/browser/view/PRJEB29739). Accessions can be found in Table S6.

## Author Statements

**Conceptualization**: David M. Aanensen, Iruka N. Okeke, Chikwe Ihekweazu; **Data curation**: Ayorinde O. Afolayan, Anthony Underwood; **Formal Analysis**: Ayorinde O. Afolayan, Anthony Underwood, Oyeniyi S. Bejide, Iruka N. Okeke; **Funding acquisition**: David M. Aanensen, Aaron O. Aboderin, Iruka N. Okeke; **Investigation**: Ayorinde O. Afolayan, Abiodun Egwuenu, Erkison Ewomazino Odih, Oyeniyi S. Bejide, Aaron O. Aboderin, Iruka N. Okeke; **Methodology**: Ayorinde O. Afolayan, Aaron O. Aboderin, Anderson O. Oaikhena, Erkison Ewomazino Odih, Oyeniyi S. Bejide, Veronica O. Ogunleye, Adeyemi T. Adeyemo, Abolaji T. Adeyemo, Anthony Underwood, Silvia Argimón, Monica Abrudan, David M. Aanensen, Iruka N. Okeke; **Project administration**: Abiodun Egwuenu, Chikwe Ihekweazu, Aaron O. Aboderin, Iruka N. Okeke; **Resources:** Anthony Underwood, Silvia Argimón, Veronica O. Ogunleye, Adeyemi T. Adeyemo, Abolaji T. Adeyemo, David M. Aanensen, Iruka N. Okeke; **Software:** Ayorinde O. Afolayan, Silvia Argimón, Anthony Underwood; **Supervision:** Aaron O. Aboderin, Anthony Underwood, Silvia Argimón, Chikwe Ihekweazu, David M. Aanensen, Iruka N. Okeke; **Validation:** All authors; **Visualization:** Ayorinde O. Afolayan, Iruka N. Okeke; **Writing – original draft:** Ayorinde O. Afolayan; **Writing – review & editing:** All authors.

## Acknowledgments/Funding

We thank Damilola Q. Olaoye, Chinenye Ekemezie, Ifeoluwa Akintayo and Ifeoluwa Komolafe for excellent technical assistance.

This work was supported by Official Development Assistance (ODA) funding from the National Institute of Health Research [grant number 16_136_111] and the Wellcome Trust grant number 206194. INO was an African Research Leader supported by the UK Medical Research Council (MRC) and the UK Department for International Development (DFID) under the MRC/DFID Concordat agreement that is also part of the EDCTP2 program supported by the European Union. The funders had no role in the content, crafting or submission of this paper.

## Ethics

Isolates were obtained as part of the surveillance efforts in line with Nigeria’s national action plan and/or for bloodstream pathogen and *Escherichia coli* research. In Nigeria. Ethical approval for research using these isolates was obtained from the University of Ibadan/University College Hospital and the Obafemi Awolowo University (OAU) Teaching Hospitals complex ethics commitees. Respective IRB registration numbers are UI/EC/15/093 and ERC/2017/05/06.

## Supplementary Tables

**Table S1:** Resistance profiles of ExPEC isolates.

| Resistance Profile (number) | Resistance Profile code |
| --- | --- |
| AMP CIP CRO CXM CXMA GEN NAL SXT (10) | RP-1 (MDR) |
| AMP CIP CXMA GEN NAL SXT (8) | RP-2 (MDR) |
| AMP CIP CRO CXM CXMA FEP GEN NAL SXT (6) | RP-3 (MDR) |
| AMP CIP NAL SXT (6) | RP-4 (MDR) |
| AMP SXT (4) | RP-5 |
| SXT (4) | RP-6 |
| AMP CIP CRO CXM CXMA FEP GEN NAL SFP SXT TZP (3) | RP-7 (MDR) |
| AMP CIP CRO CXM CXMA FEP NAL SXT (3) | RP-8 (MDR) |
| AMP CIP CRO CXM CXMA NAL SXT (3) | RP-9 (MDR) |
| AMP NAL SXT (3) | RP-10 (MDR) |
| AMP CIP CRO CXM CXMA FEP GEN NAL NIT SXT (2) | RP-11 (MDR) |
| AMP CIP CRO CXM CXMA FEP GEN NAL SXT TZP (2) | RP-12 (MDR) |
| AMP CIP CXM CXMA NAL SXT TZP (2) | RP-13 (MDR) |
| AMP CIP CXMA GEN NAL NIT SXT (2) | RP-14 (MDR) |
| AMP CIP CRO CXM CXMA GEN NAL SXT TZP (1) | RP-other (MDR) |
| AMP CIP CRO CXM CXMA SXT (1) | RP-other (MDR) |
| AMP CIP CXM CXMA GEN NAL SXT TZP (1) | RP-other (MDR) |
| AMP CIP CXMA NAL SXT (1) | RP-other (MDR) |
| AMP CIP CXMA NAL SXT TZP (1) | RP-other (MDR) |
| AMP CIP GEN NAL SXT (1) | RP-other (MDR) |
| AMP CXMA SXT (1) | RP-other (MDR) |
| AMP GEN NAL SXT TZP (1) | RP-other (MDR) |
| NAL SXT TZP (1) | RP-other (MDR) |

**Table S2:** Summary of Antibiotic Susceptibility Test data showing the number of *E. coli* isolates that were resistant to the named antibiotics.

| Drug class | Drug | Resistance % (resistance (n)/species (n)) |
| --- | --- | --- |
| Aminoglycosides | Gentamicin (GEN) | 55.20% (37/67) |
| Beta-lactams/penicillins | Ampicillin (AMP) | 92.50% (62/67) |
|  | Piperacillin/tazobactam (TZP) | 17.90% (12/67) |
| Cephalosporins (2nd gen.) | Cefuroxime (CXM) | 50.70% (34/67) |
|  | Cefuroxime Axetil (CXMA) | 70.10% (47/67) |
| Cephalosporins (3rd gen.) | Cefoperazone.Sulbactam (SFP) | 4.50% (3/67) |
|  | Ceftriaxone (CRO) | 46.30% (31/67) |
| Cephalosporins (4th gen.) | Cefepime (FEP) | 23.90% (16/67) |
| Other antibacterials | Nitrofurantoin (NIT) | 6.00% (4/67) |
| Quinolones | Ciprofloxacin (CIP) | 79.10% (53/67) |
|  | Nalidixic acid (NAL) | 85.10% (57/67) |
| Trimethoprim | Trimethoprim/sulfamethoxazole (SXT) | 100.0% (67/67) |

**Table S3:** Degree of Agreement between phenotypic antimicrobial susceptibility tests and genotypic predicted antimicrobial resistance.

| drug | Tests (N) | concordance (%) | TP | FP | TN | FN | specificity | Specificity lower limits | Specificity upper limits | sensitivity | Sensitivity lower limits | Sensitivity upper limits |
| --- | --- | --- | --- | --- | --- | --- | --- | --- | --- | --- | --- | --- |
| Aminoglycosides | 67 | 55.22 | 37 | 30 | 0 | 0 | 0 | 0 | 0.115703308 | 1 | 0.905109413 | 1 |
| Beta-lactams | 67 | 71.64 | 45 | 1 | 3 | 18 | 0.75 | 0.19412045 | 0.993690537 | 0.714285714 | 0.586538974 | 0.821070793 |
| Cephalosporins | 67 | 68.66 | 46 | 20 | 0 | 1 | 0 | 0 | 0.168433471 | 0.978723404 | 0.887062283 | 0.999461468 |
| Quinolones | 67 | 86.57 | 56 | 7 | 2 | 2 | 0.222222222 | 0.028144973 | 0.600093574 | 0.965517241 | 0.880922689 | 0.995796353 |
| Trimethoprim | 67 | 100 | 67 | 0 | 0 | 0 | NA | 0 | 1 | 1 | 0.946430346 | 1 |

**Table S3b.**
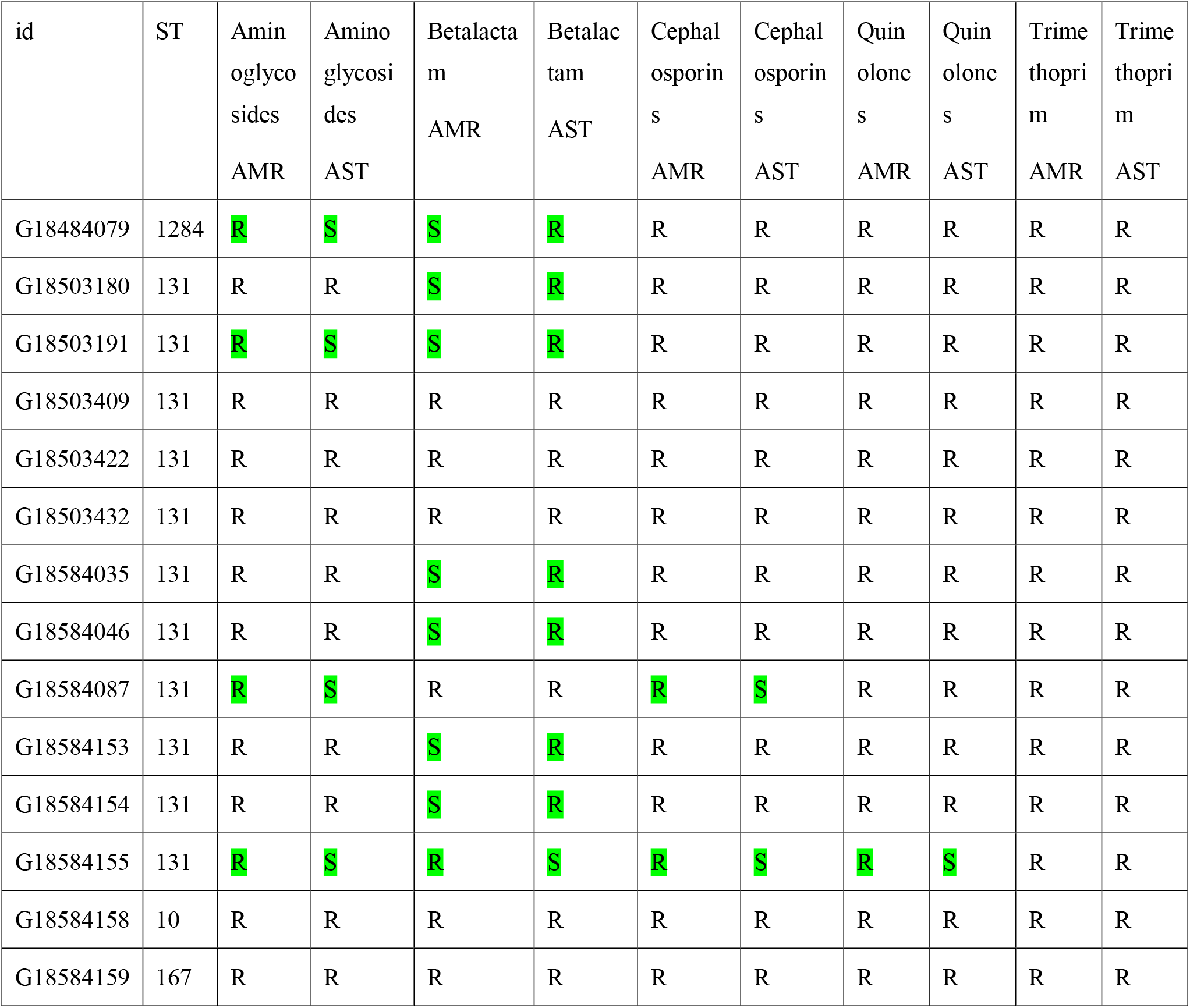
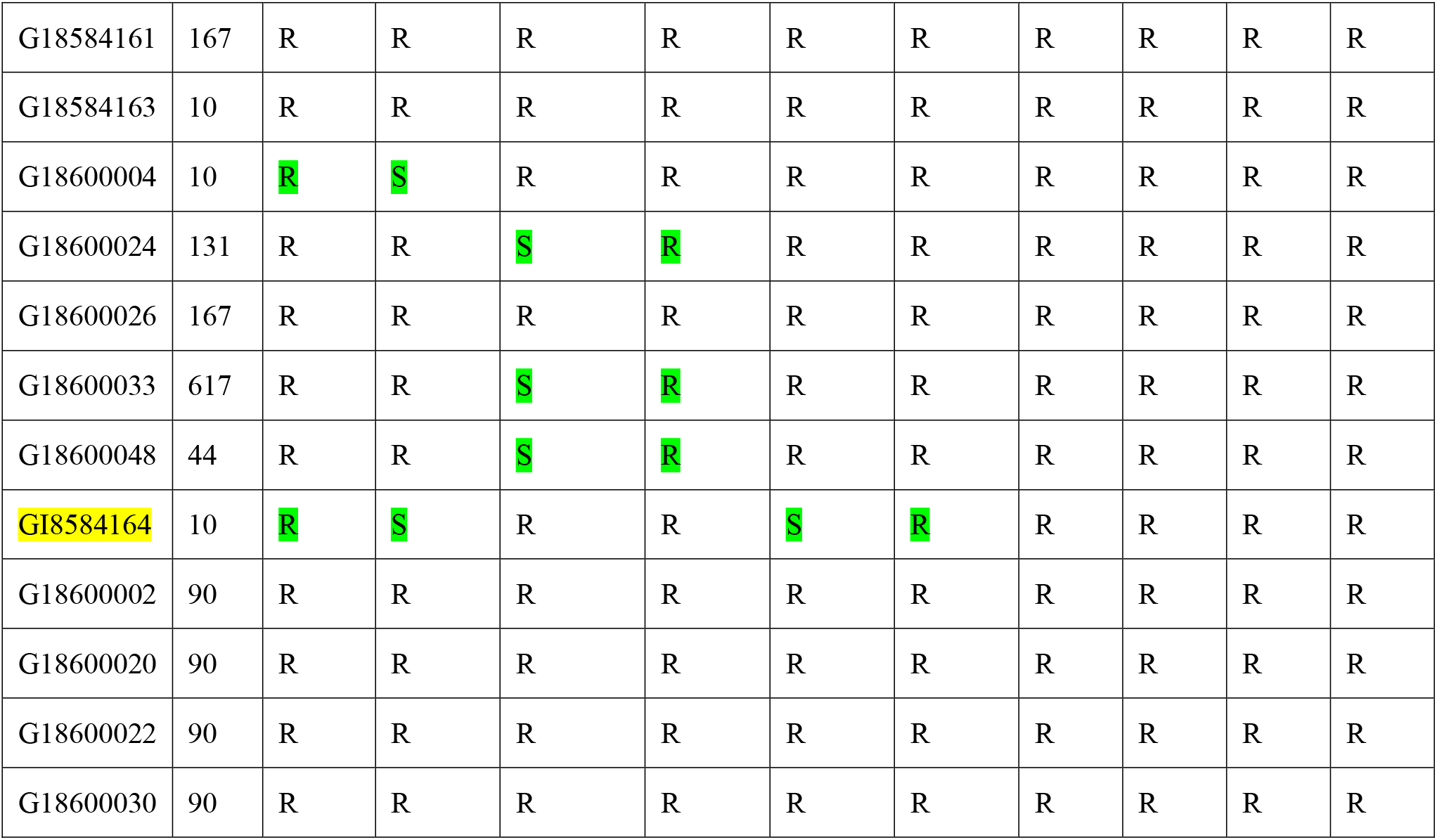
Degree of Agreement between phenotypic antimicrobial susceptibility tests and genotypic predicted antimicrobial resistance in isolates belonging to the ST131, ST90, and ST10-167 clades.

| id | ST | Amin<br>oglyco<br>sides<br>AMR | Amino<br>glycosi<br>des<br>AST | Betalacta<br>m<br>AMR | Betalac<br>tam<br>AST | Cephal<br>osporin<br>s<br>AMR | Cephal<br>osporin<br>s<br>AST | Quin<br>olone<br>s<br>AMR | Quin<br>olone<br>s<br>AST | Trime<br>thopri<br>m<br>AMR | Trime<br>thopri<br>m<br>AST |
| --- | --- | --- | --- | --- | --- | --- | --- | --- | --- | --- | --- |
| G18484079 | 1284 | R | S | S | R | R | R | R | R | R | R |
| G18503180 | 131 | R | R | S | R | R | R | R | R | R | R |
| G18503191 | 131 | R | S | S | R | R | R | R | R | R | R |
| G18503409 | 131 | R | R | R | R | R | R | R | R | R | R |
| G18503422 | 131 | R | R | R | R | R | R | R | R | R | R |
| G18503432 | 131 | R | R | R | R | R | R | R | R | R | R |
| G18584035 | 131 | R | R | S | R | R | R | R | R | R | R |
| G18584046 | 131 | R | R | S | R | R | R | R | R | R | R |
| G18584087 | 131 | R | S | R | R | R | S | R | R | R | R |
| G18584153 | 131 | R | R | S | R | R | R | R | R | R | R |
| G18584154 | 131 | R | R | S | R | R | R | R | R | R | R |
| G18584155 | 131 | R | S | R | S | R | S | R | S | R | R |
| G18584158 | 10 | R | R | R | R | R | R | R | R | R | R |
| G18584159 | 167 | R | R | R | R | R | R | R | R | R | R |
| G18584161 | 167 | R | R | R | R | R | R | R | R | R | R |
| G18584163 | 10 | R | R | R | R | R | R | R | R | R | R |
| G18600004 | 10 | R | S | R | R | R | R | R | R | R | R |
| G18600024 | 131 | R | R | S | R | R | R | R | R | R | R |
| G18600026 | 167 | R | R | R | R | R | R | R | R | R | R |
| G18600033 | 617 | R | R | S | R | R | R | R | R | R | R |
| G18600048 | 44 | R | R | S | R | R | R | R | R | R | R |
| GI8584164 | 10 | R | S | R | R | S | R | R | R | R | R |
| G18600002 | 90 | R | R | R | R | R | R | R | R | R | R |
| G18600020 | 90 | R | R | R | R | R | R | R | R | R | R |
| G18600022 | 90 | R | R | R | R | R | R | R | R | R | R |
| G18600030 | 90 | R | R | R | R | R | R | R | R | R | R |

**Table S3c:** List of genes predicted to confer resistance to antibiotics belonging to the drug classes in Table S3a

| **Drug Classes** | **AMR genes** |
| --- | --- |
| Aminoglycosides | aac(6')-Ie/aph(2'')-Ia, aac(6')-Ib-cr5, catB8/aac(6')-Ib', aac(6')-30/aac(6')-Ib', aac(3)-IIe, aac(3)-IId, aac(6')-Ib4, ant(2'')-Ia, aph(3')-Ia, aadA2, aph(3'')-Ib, aph(6)-Id, aadA5, aadA13, aadA1, aadA8 |
| Beta-lactams | blaTEM-1, blaOXA-2, blaTEM-90, blaTEM-57, blaTEM-40, blaTEM-84, blaZ, blaR1, blaTEM-135 |
| Cephalosporins | blaCTX-M-15, blaEC-15, blaEC-5, blaOXA-1, blaEC-16, blaEC-18, blaCTX-M-27, blaEC-8, blaCMY-2, blaEC-19, blaCMY-42, blaTEM-187, blaVEB-1 |
| Quinolones | gyrA, parC, and pare mutations, qepA4, qnrS1, qepA8, qnrVC4 |
| Trimethoprim | dfrA12, dfrA17, dfrA7, dfrA14, dfrA29, dfrF, dfrB4, dfrA5, dfrA1 |

**Table S4:**
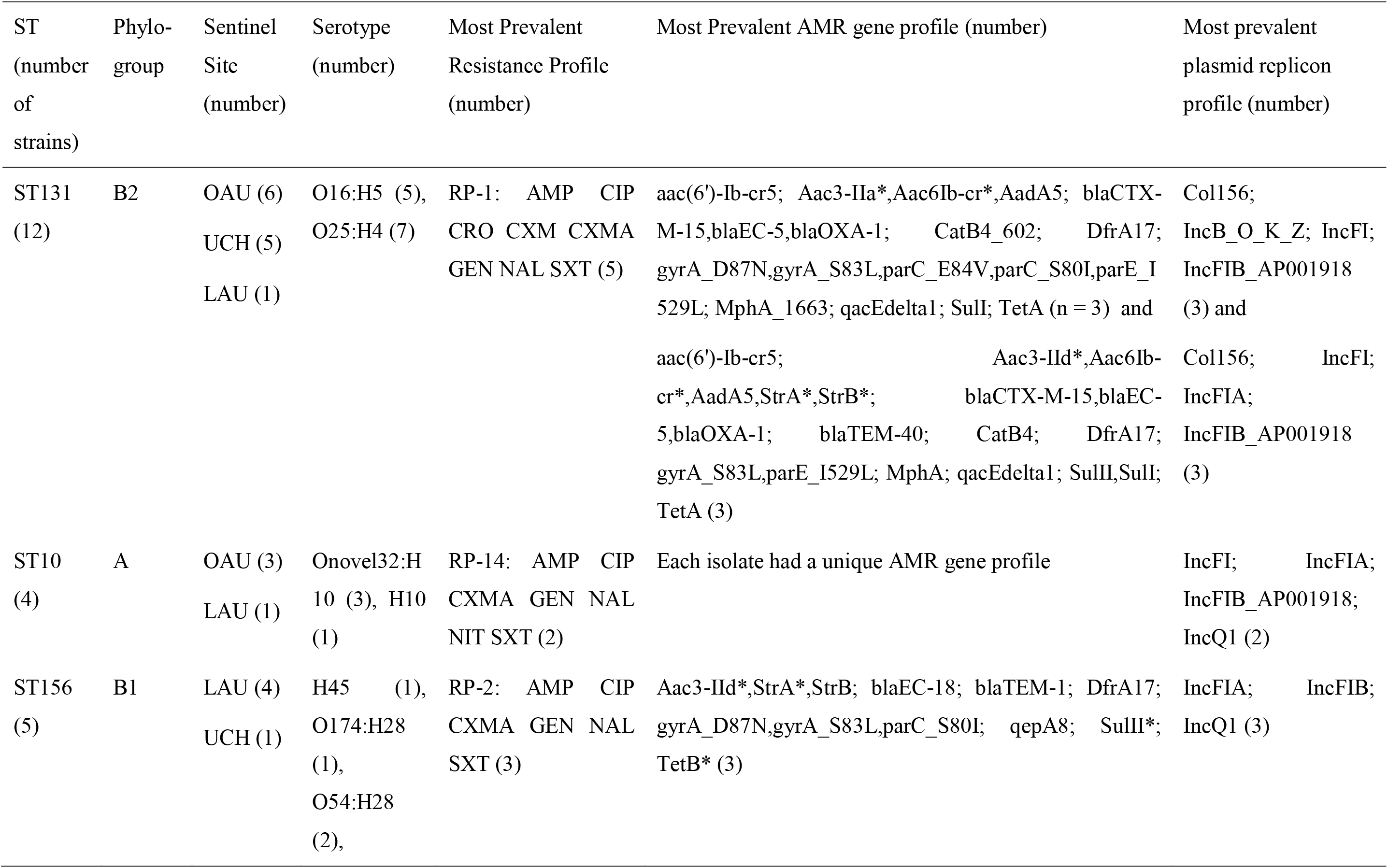
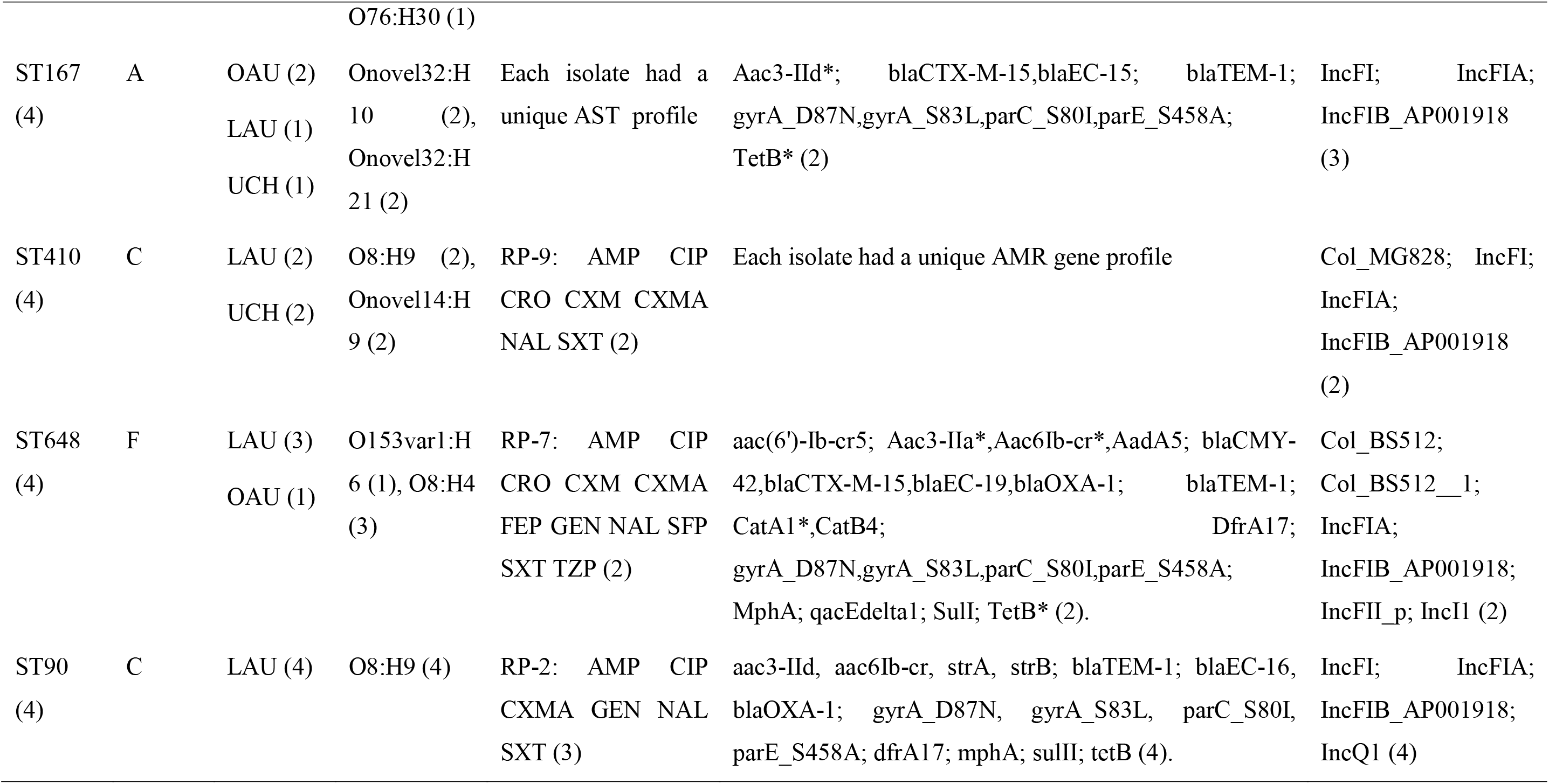
Genotypic characteristics of the most common STs in the invasive microbial population.

**Table S5:**
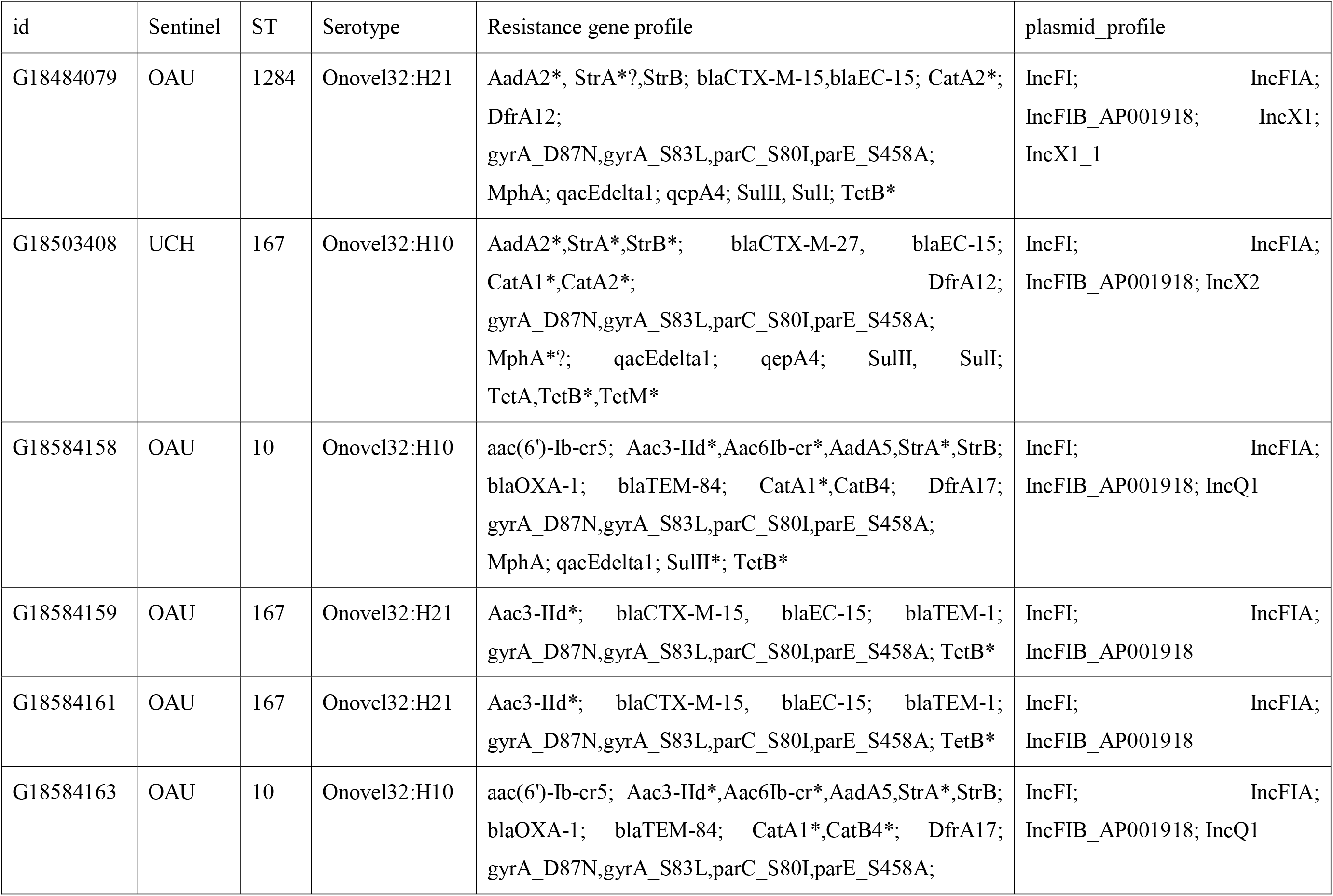
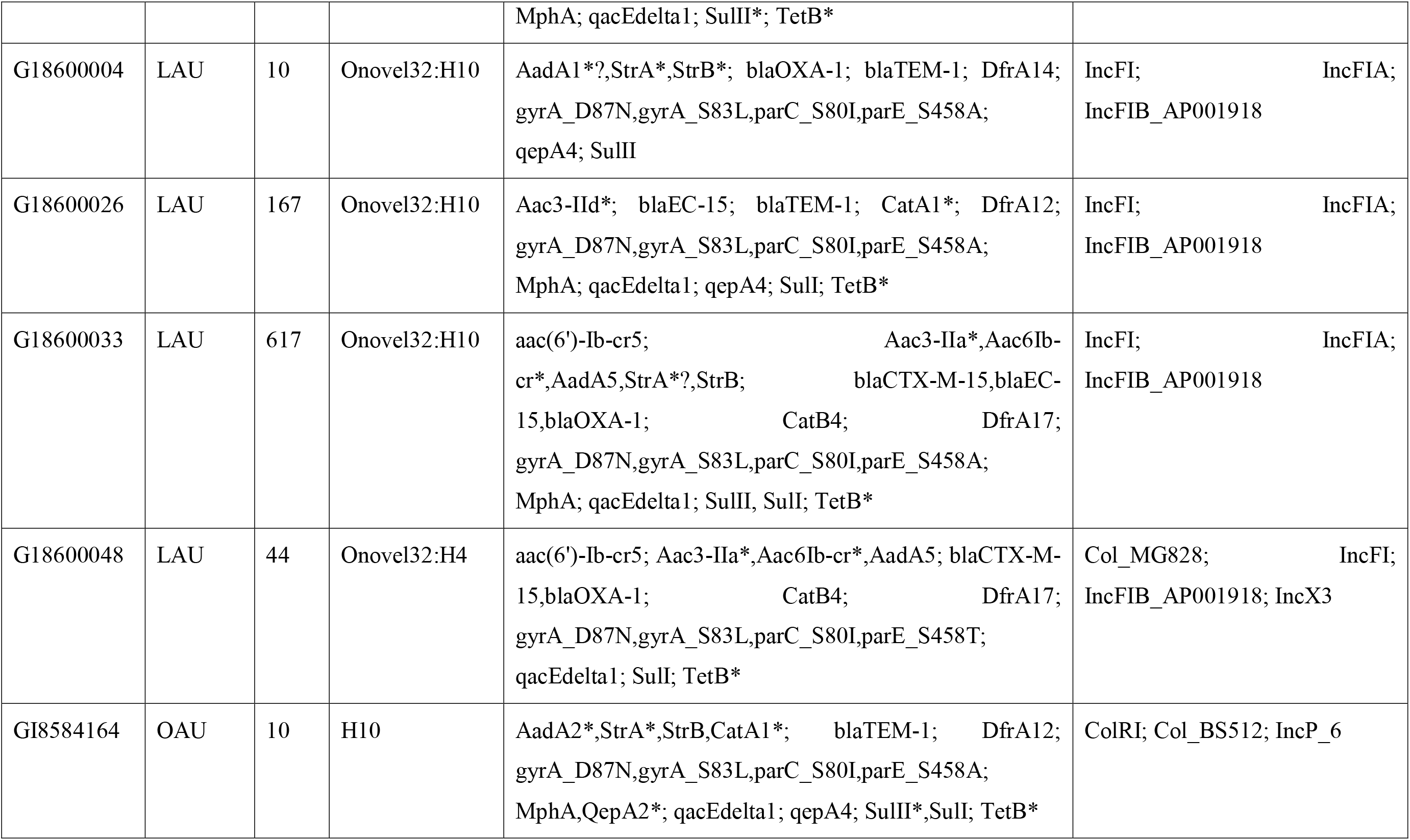
Genotypic characteristics of Onovel32 genomes in the invasive microbial population.

**Supplementary Figure 1.**
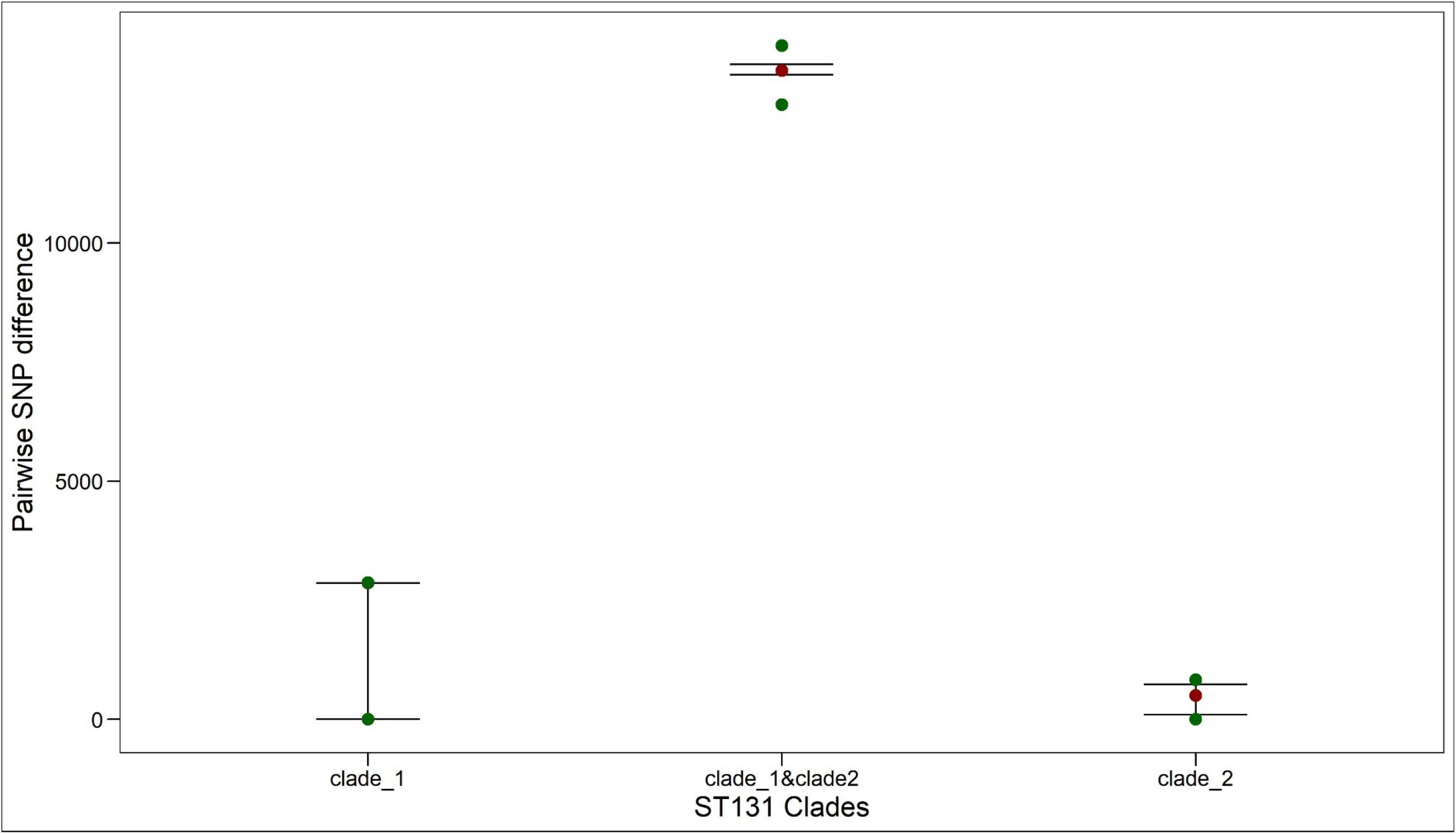
Interclade (ST131 clade 1 and ST131 clade 2) distance between lineage ST131 isolates is greater than 10000 SNPs. The “whisker” plot shows the 25^th^ percentile (lower point; green), median (red point), and the 75^th^ percentile (upper point; green) of the pairwise SNP distances within and between lineages.

**Supplementary Figure. 2:**
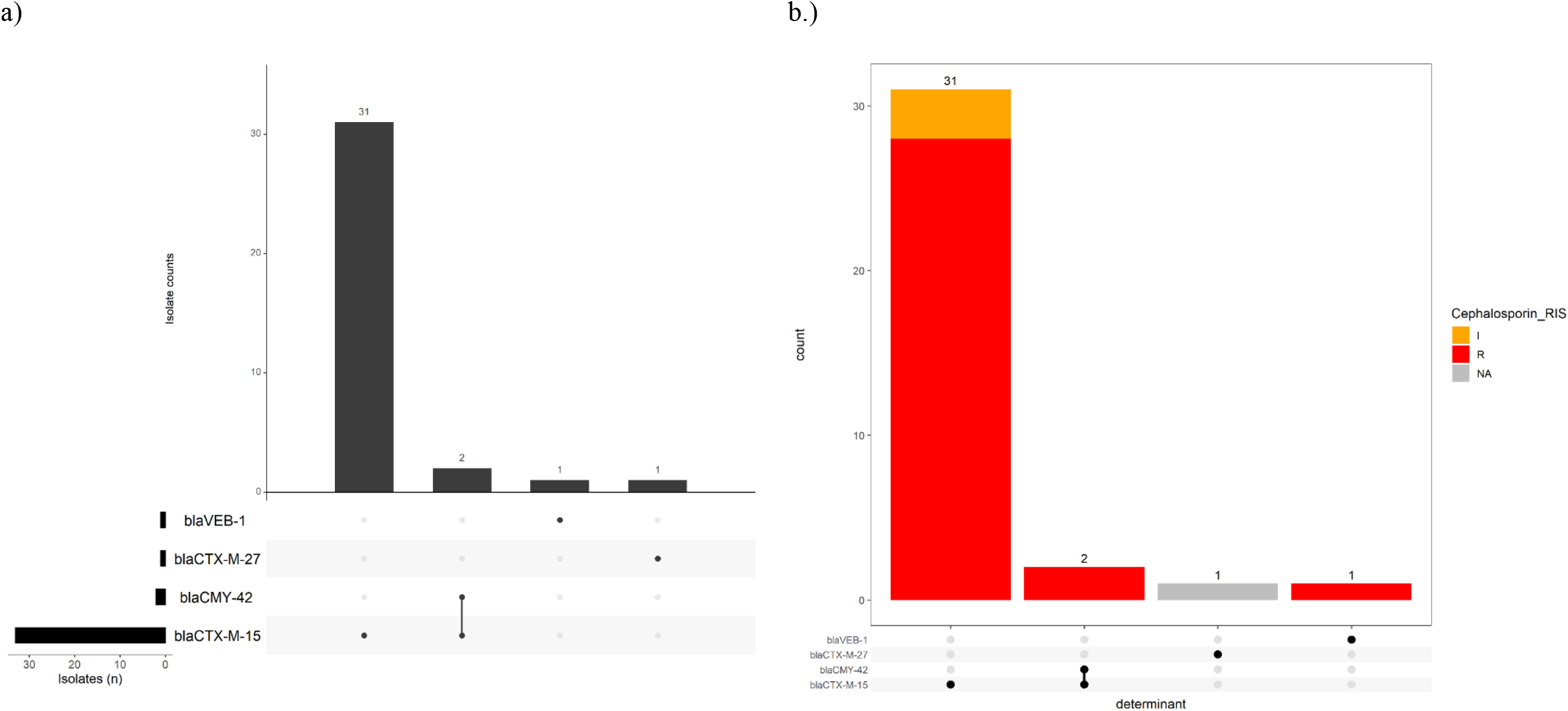
ESBL genes in ExPEC isolates. The upset plots demonstrate (a, b) the number of ExPEC isolates carrying each combination of genes conferring resistance to cephalosporins and (b), is coloured by the proportion of observed phenotypic antimicrobial susceptibility, and is ordered in descending order by the frequency of resistance gene profiles observed. The side bar chart (a) demonstrates the number of isolates that carry each of the resistance genes. The dots and lines between dots at the base of the main bar chart (and the right side of the side bar chart) show the co-resistance gene profile of the ExPEC isolates. From the legend of the coloured upset plot, NA means that there is no available data on phenotypic antimicrobial susceptibility data for the isolate.

**Supplementary Figure 3:**
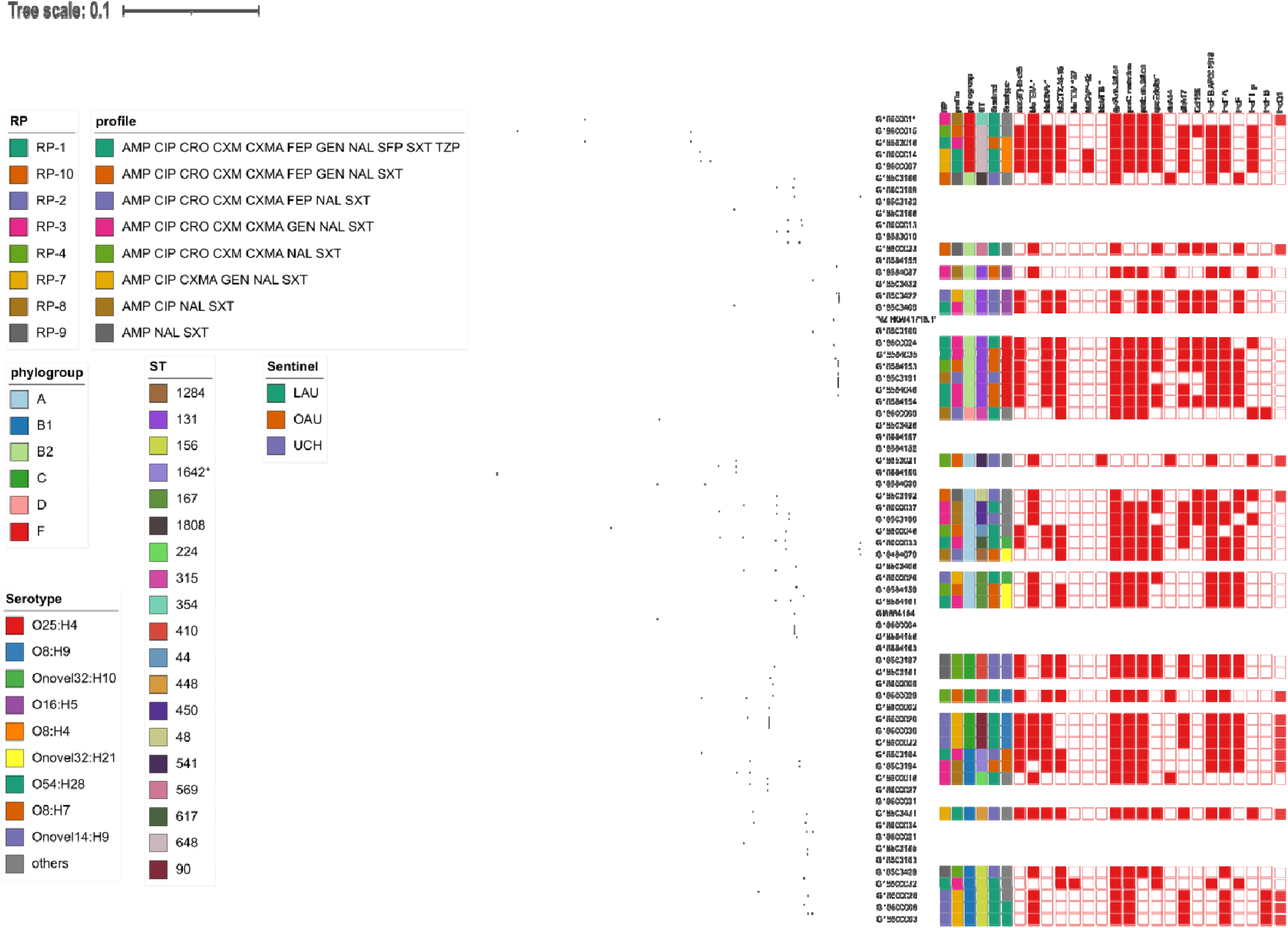
Maximum likelihood SNP tree of bloodstream *E. coli* isolates sequenced for this study. Metadata for the 8 most common resistance profiles are shown in this figure. For the AMR genes and plasmid replicons, red colour indicates presence of the gene.

